# Cost-effectiveness of transplanting older candidates with acceptable quality deceased donor kidneys

**DOI:** 10.1101/2025.06.04.25328892

**Authors:** Matthew B Kaufmann, Jane C Tan, Douglas K Owens, Glenn M Chertow, Jeremy D Goldhaber-Fiebert

## Abstract

**Importance:** Many acceptable quality deceased donor kidneys go unused every year. Older transplant candidates are more vulnerable to rapid health decline.

**Objective:** To assess the cost-effectiveness of increasing the rate of kidney transplantation in older patients with end-stage kidney disease by using acceptable quality deceased donor kidneys.

**Design:** This cost-effectiveness analysis utilizes a microsimulation model of the kidney transplantation process for older adult candidates over a lifetime horizon.

**Setting:** Health state transition probabilities are derived from Scientific Registry of Transplant Recipient data. Costs and quality-of-life weights are derived from published literature and United States Renal Data System annual reports, all of which are varied in a probabilistic sensitivity analysis.

**Participants:** A synthetic population of deceased donor kidney transplant candidates 65 or older.

**Interventions:** Increasing the rate of transplantation in 5% increments higher than the status quo rate from 5% to 25% using acceptable quality deceased donor kidneys.

**Main Outcomes and Measures:** The primary outcomes are the number of key waitlist and post-transplant outcomes, costs, quality-adjusted life-years (QALYs), incremental cost-effectiveness ratios (ICERs).

**Results:** We estimated there would be 141 fewer waitlist deaths per 10,000 candidates if the rate of deceased donor transplantation were increased by 25% from the status quo rate. Increasing the rate of deceased donor transplantation by 25% costs $8,100 per QALY gained or is cost-saving from the healthcare sector and modified healthcare sector perspectives, respectively. From the healthcare sector perspective, a 25% increase in the rate of deceased donor transplantation is the preferred strategy in all probabilistic sensitivity analysis samples for willingness-to-pay thresholds ≥$40,000 per QALY gained.

**Conclusions and Relevance:** Increasing the rate of kidney transplantation in older adults, even using acceptable quality deceased donor kidneys, would be cost-effective or cost-saving.

## Introduction

The United States (US) kidney transplant system does not operate at optimal efficiency. There were 71,523 patients on the kidney transplant waitlist at the end of 2022 and of those who were listed 5 years ago, 48.7% had received a transplant.^1^ For many patients, longevity and quality-of-life would be enhanced by kidney transplantation in lieu of dialysis, a treatment that sustains life, but rarely restores health. Despite the urgent need, roughly one in five recovered deceased donor kidneys were not utilized.^2^ The proportion of recovered but ultimately unused deceased donor kidneys has steadily risen, from 5.1% in 1988 to 24.6% in 2021.^2,3^ An estimated 17,435 (62%) unused donor kidneys in the United States from 2004 to 2014 would have been used in France’s more liberal organ allocation system.^4^ The French system has many differences from the US system including less regulatory scrutiny and a prohibition on performing a biopsy on deceased donor kidneys before transplantation.^4^ Biopsies are prohibited because they are believed to add little information about the quality of donor’s kidney while prolonging the time-sensitive allocation process.^5^

Older patients with end-stage kidney disease (ESKD) are transplanted at sharply lower rates than their younger counterparts.^6^ The probability of being evaluated, listed, and receiving a transplant declines as patients age and accumulate comorbid conditions and complications associated with ESKD.^7^ In 2021, those aged 65 or older comprised 42.8% of all patients with ESKD but only comprised 23.6% of transplant candidates, and 20.6% of patients who received a kidney transplant. Older transplant candidates receiving dialysis might be willing to trade off receiving a “lower quality” deceased donor kidney (e.g., a kidney from an older donor, or a donor with a history of hypertension) for a shorter waiting time.^8^ While older candidates may have heterogeneous preferences, the current system makes it difficult to fully incorporate patient preferences into decision-making. Thus, systematically operationalizing patient choice into critical moments of decision-making in the modern-day clinical platform is exceedingly challenging The intense focus on transplant performance metrics has fostered increasingly conservative practices at many transplant centers. Policy metrics for evaluating transplant centers have been at odds with increasing access to transplantation. For example, transplant centers may have reservation over a disproportionate pool of higher risk (including older) candidates and concern over non-judicious use of lower quality deceased kidneys for worry over negatively affecting results and potentially threatening the viability of the transplant program itself.^9^ Metrics that include first-year patient and graft survival, waitlist mortality rates, and transplantation rates bias against transplantation of older and sicker patients. Transplant centers should be able to provide health-enhancing, high-quality, high-value care for all patients, not only those in the best of health with the lowest rates of complications.

We hypothesized that more widespread use of deceased donor kidneys of lower but acceptable quality would improve outcomes for older patients with ESKD and would prove to be cost-effective or cost-saving. In support of this policy discussion, we performed a model-based cost-effectiveness analysis of evaluating the benefits, harms, effectiveness, and cost-effectiveness of increasing the rate of transplantation among older candidates by using lower but acceptable quality recovered deceased donor kidneys.

## Methods

### Model Description

We used a previously developed discrete time microsimulation model of the deceased donor kidney transplant process for older adults in the United States developed by the authors of this study.^10^ The model follows patients on the waitlist, tracking outcomes including deceased donor transplantation, living donor transplant, waitlist mortality, and other waitlist removals. Other waitlist removals encompass removals for any reason other than death or transplantation, including removals due to declining health. For those who receive a deceased donor kidney, the model then tracks 30-day post-transplant, mutually exclusive outcomes: 1) no adverse events, 2) delayed graft function (DGF), 3) graft loss, or 4) death. We do not differentiate between graft loss and acute rejection as acute rejection is a rare cause of graft loss in this population. Depending on their graft function and survival after the first 30 days, the model follows simulated patients over their remaining lives tracking events including graft failure and death (**Figure 1)**. We assumed that there is no chance for re-transplantation.

**Figure 1:**
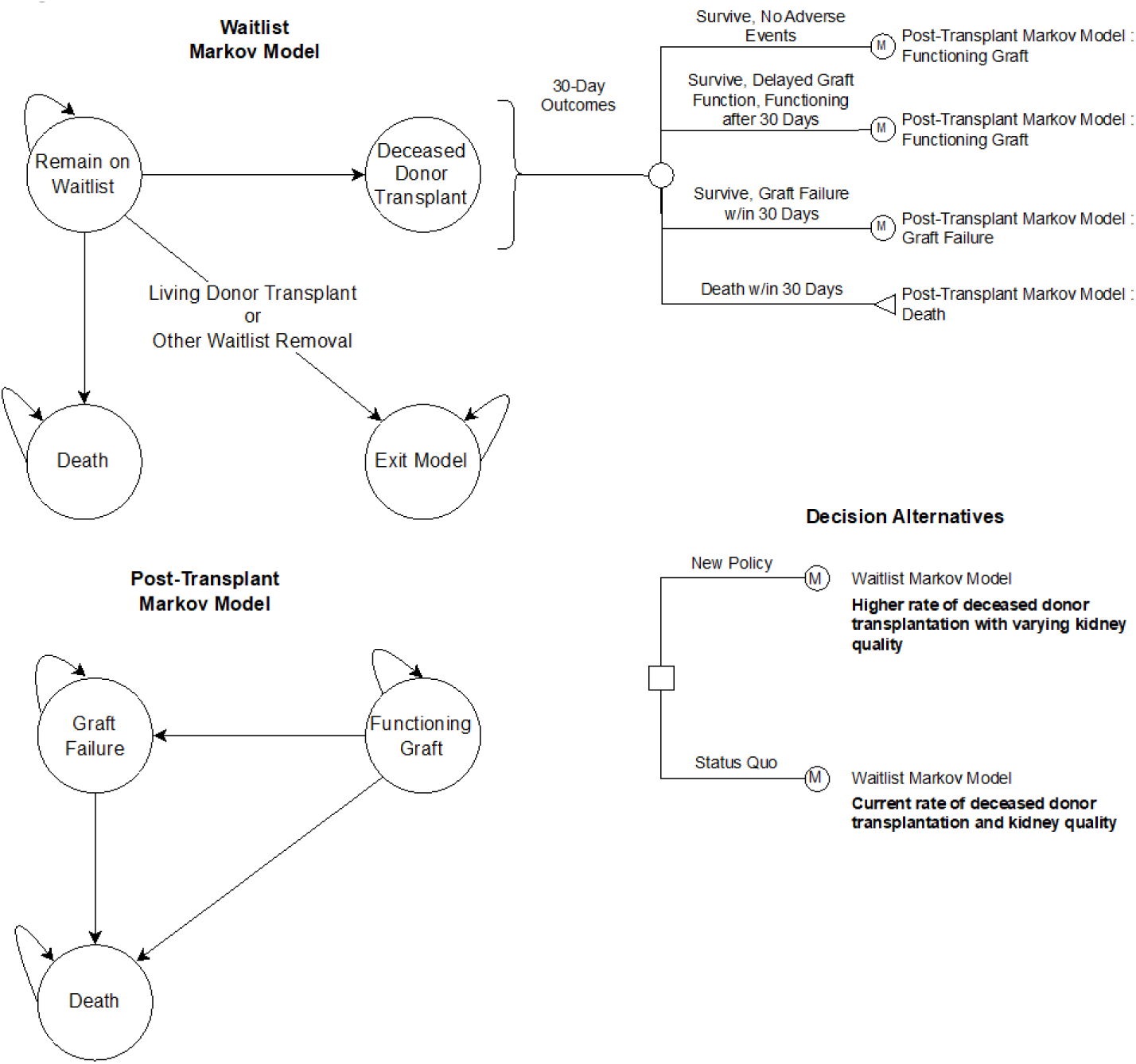
Model Diagram. Reprinted from “Deceased donor kidney transplantation for older transplant candidates – a new microsimulation model for determining risks and benefits,” by M.B. Kaufmann, 2023, *Medical Decision Making, 43*(5), p. 580. Copyright 2023 by The Authors. Reprinted with permission.

### Data and Sources

Transitions between health states depend on a series of risk equations that involve patient and donor characteristics. In our previous work, we estimated risk equations from Scientific Registry of Transplant Recipient data which represents all candidates and recipients who were waitlisted and transplanted between 2010-2019.^11,12^ To account for competing risks, we employed likelihood-based model calibration to simultaneously match a set of waitlist and transplant outcomes (details in Supplement, Methods S3). For the present cost-effectiveness analysis, we updated the calibration using 64 equally weighted sets of calibrated parameters, all of which were consistent with calibration targets (see Kaufmann et al., 2023 for values of parameters and equation forms).^10^

Kidney transplant care costs were included in the analysis conducted from the healthcare sector perspective, which encompassed dialysis, transplant, and post-transplant events and complications. We also included Organ Acquisition Costs Center (OACC) payments, which are sizeable payments to transplant programs to reimburse the costs for transplant evaluation and waitlist management.^13^ Patient and caregiver time (opportunity) costs were added to our analysis from the modified healthcare sector perspective. We calculated caregiver time costs as the average number of hours per month spent caregiving multiplied by mean wage estimates from the Bureau of Labor Statistics (BLS).^14–16^ We assumed that all transplant candidates have a caregiver. We calculated patient time costs as the average number of hours per month a patient receives care multiplied by the same BLS estimates of mean wages. The number of hours of caregiving and receiving care were generated through discussions with experts in transplant nephrology (**Table 1**). We assumed that older transplant candidates have the same time costs as a younger working-age population. Excluding these costs would imply that their time no longer has value when they are not working.

**Table 1:**
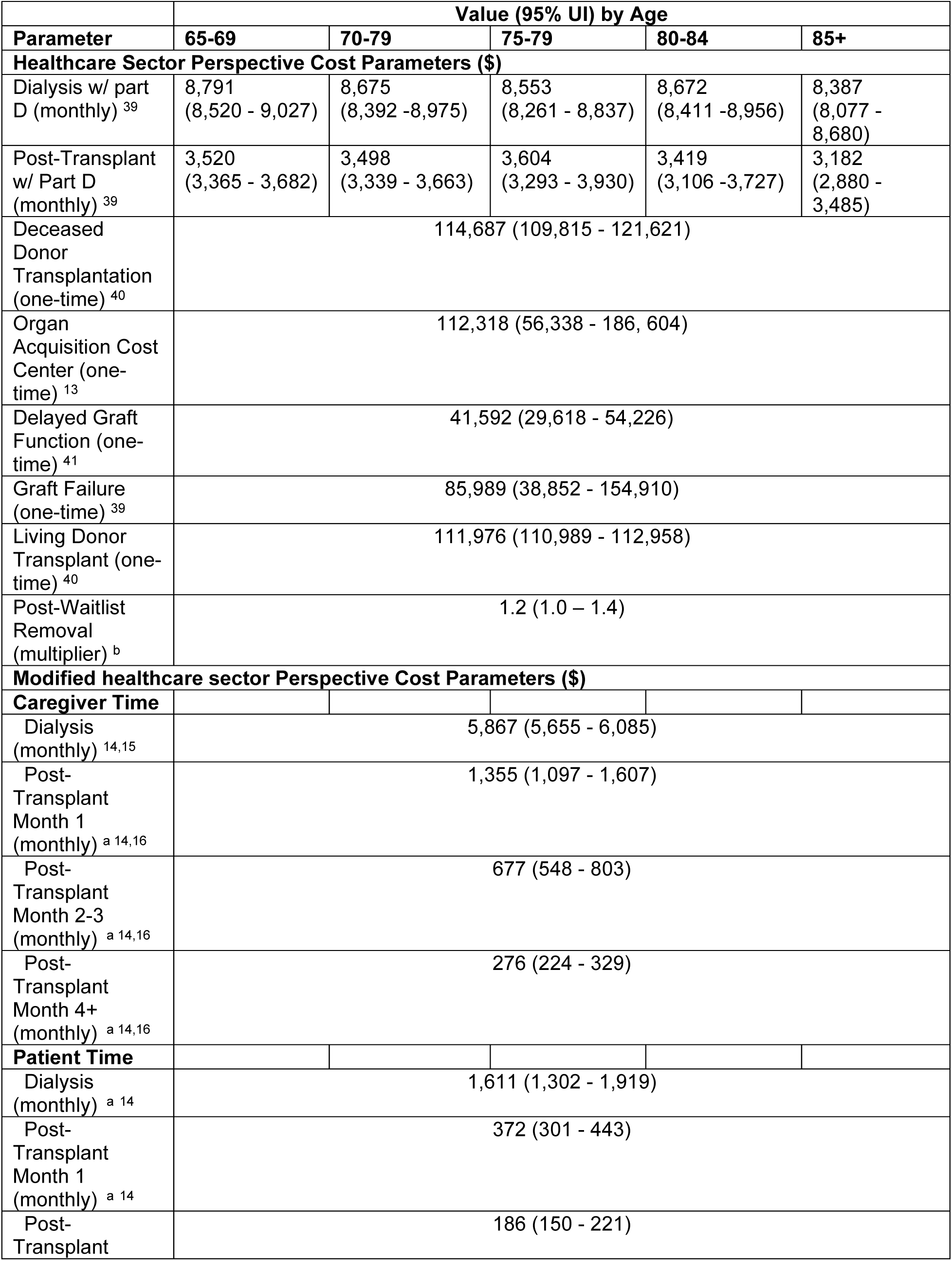
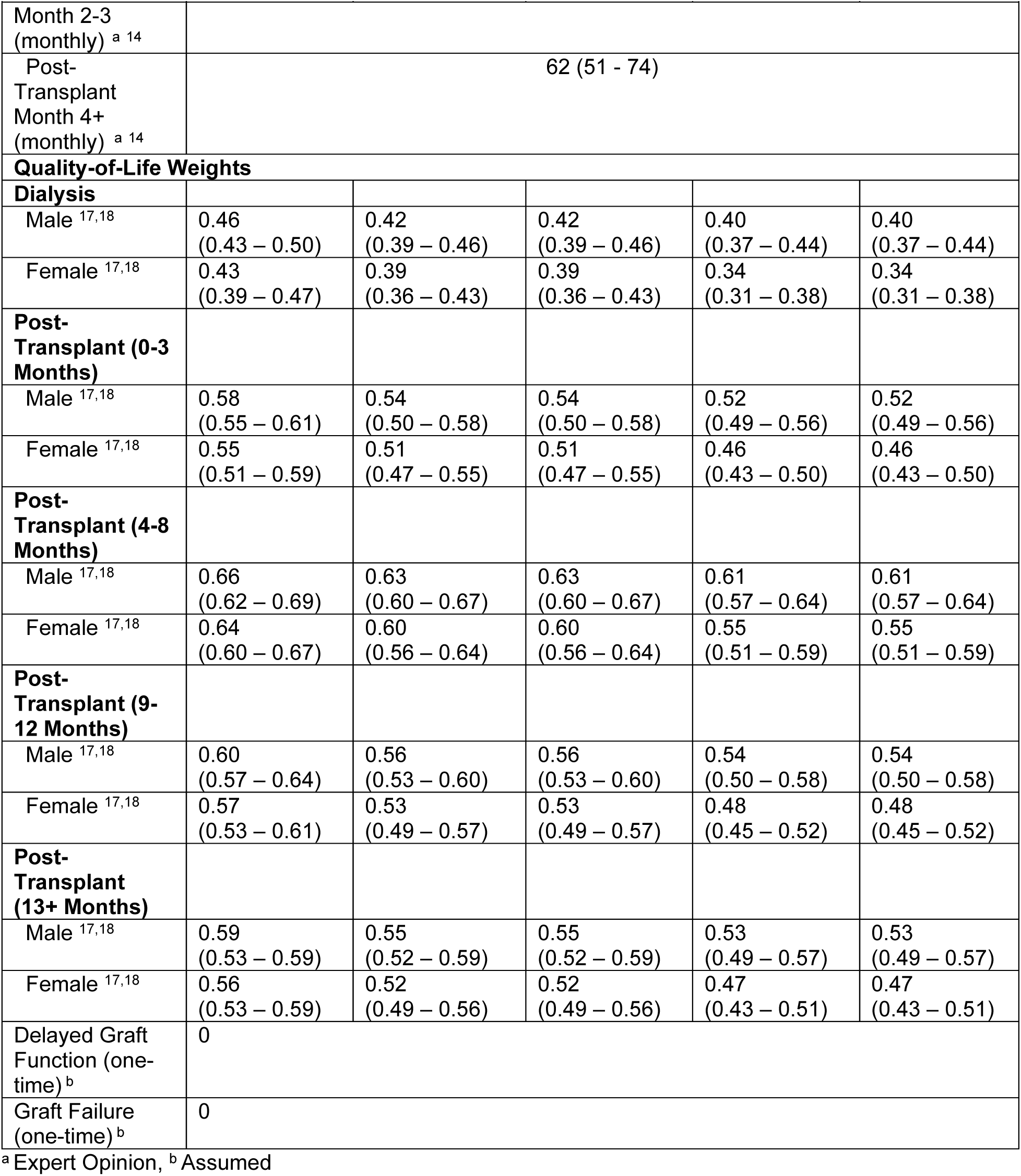
Model Inputs with Uncertainty Intervals.

We assumed that patients on the waitlist received dialysis and that patients who were removed from the waitlist before transplantation or death incurred additional healthcare costs beyond the average costs for patients on dialysis. This assumption is made because a common reason for removal other than for death or transplantation is because of declining health that would make them unable to receive a transplant. We assumed a 20% increase in dialysis costs for patients removed from the waitlist and varied the increment in our sensitivity analysis.

We derived health-related quality-of-life weights that are age-and sex-specific pre-transplant as well as age-, sex-, and time since transplant-specific post-transplant (**Table 1**).^17,18^ Additional information on our health-related quality-of-life calculations can be found in the Supplement, Methods S2. We also conservatively assumed that during graft failure (and prior to graft function for those who experienced DGF), a patient’s health-related quality-of-life was 0. In our probabilistic sensitivity analysis, we assumed a rank ordering where health-related quality-of-life weights decrease with age. The same applies to health-related quality-of-life weights by time since transplantation. We used the method outlined in Goldhaber-Fiebert & Jalal, 2016 to preserve this rank order.^19^

### Interventions

To determine the effects of hypothetical policies, we simulated an increased rate of transplantation from current practice with a shift in the distribution of kidney quality due to the utilization of lower but acceptable quality deceased donor kidneys. The increased rate of transplantation will likely depend on how transplant centers respond to policies that make use of additional acceptable quality kidneys, which is currently unknown. Hence, we considered the status quo (0% increase in the transplantation rate) along with 5 potential policies that would increase the transplantation rate by 5%, 10%, 15%, 20%, and 25% respectively to understand the health and economic implications of a range of hypothetical policies. We assumed that with the implementation of policies to increase the transplantation rate for older transplant candidates, the average quality of deceased donor kidneys declines such that it closely resembles the distribution of kidney quality as measured by kidney donor profile index (KDPI) of kidneys utilized in France’s system. KDPI is meant to reflect the risk of graft loss, with higher values reflecting higher risk. It is also associated with a higher probability of DGF.^20^ Thus, the distribution of KDPI will now reflect a higher utilization of high KDPI donor kidneys. It is likely that a currently utilized kidney of a given KDPI is of higher quality than an unused kidney of the same KDPI. The rationale being that there are other characteristics that are not captured in KDPI that can contribute to the quality of the donor kidney. Because we used KDPI as our metric for donor kidney quality, we reflect that some proportion of newly transplanted kidneys will be lower quality by increasing their KDPI by 5 KDPI percentage points for 5% of donor kidneys. We conducted scenario analyses around the proportion of donor kidneys for which we increased the KDPI in 5% increments for up to 20% of donor kidneys.

### Analysis

Our primary outcomes include discounted lifetime costs, quality-adjusted life years (QALYs), and incremental cost-effectiveness ratios (ICERs). We also report rates of key waitlist and post-transplant outcomes. We performed two analyses, one from the healthcare sector perspective and one from the modified healthcare sector perspective in which we include patient time costs and caregiving costs. We conducted the analysis over a lifetime horizon with both future costs and QALYs discounted at 3% annually.^21^ To express all costs in 2023 USD, we used the Personal Consumption Expenditure Health Total deflator for all costs except for wage estimates for which we used the Consumer Price Index.^22,23^ We evaluated the cost-effectiveness of interventions using a $100,000 per QALY gained willingness-to-pay (WTP) threshold.^24^ We present results for the entire older adult transplant candidate population as well as by key subgroups including age, designated race/ethnicity, and diabetes status.

We explored uncertainty in our model using probabilistic sensitivity analysis (PSA). We visualized and quantified the optimal strategy at various WTP thresholds using cost-effectiveness acceptability curves (CEAC) and cost-effectiveness acceptability frontier (CEAF). We also used expected loss curves (ELC) to show the forgone benefits that would be expected to occur from choosing a suboptimal strategy depending on the WTP threshold.^25^ Additional details on the PSA can be found in the supplemental materials.

## Results

### Base Case Results

Increasing the rate of deceased donor transplantation substantially decreases the number of deaths on the waitlist and waitlist removals while increasing the total net number of people transplanted (**Table 2**). Using lower quality donor kidneys increases the risk of DGF, but shorter wait times decrease this risk. At a 5% increase in the rate of transplantation, the rate of DGF rises from the status quo rate, likely due to a small reduction in waiting time. However, as we increase the rate of deceased donor transplantation up to 25%, the rate of DGF moves back towards the status quo rate because of the offsetting risk factors. This pattern also occurs for the rate of graft failure among recipients. While lower kidney quality was associated with a higher risk of graft failure, shorter pre-transplant dialysis time had a “protective” effect.

**Table 2:** Event counts (and range) in status quo and incremental difference (and range) by scenario.

| Event | Status Quo | Incremental Difference in Event Counts by<br>Percentage Increase in Rate of Deceased Donor Transplantation |  |  |  |  |
| --- | --- | --- | --- | --- | --- | --- |
|  |  | 5% | 10% | 15% | 20% | 25% |
| Deceased Donor Transplant <sup>a</sup> | 3,623<br>(3,265 – 3,898) | 123<br>(114 – 130) | 241<br>(226 – 252) | 356<br>(338 – 368) | 468<br>(444 – 486) | 576<br>(546 – 598) |
| Living Donor Transplant <sup>a</sup> | 1,330<br>(1,162 – 1,542) | -13<br>(-16 – -8) | -25<br>(-29 – -20) | -37<br>(-42 – -31) | -49<br>(-55 – -42) | -60<br>(-69 – -52) |
| Waitlist Deaths <sup>a</sup> | 1,640<br>(1,452 – 1,799) | -30<br>(-35 – -26) | -59<br>(-67 – -50) | -87<br>(-100 – -72) | -114<br>(-129 – -98) | -141<br>(-161 – -118) |
| Other Waitlist Removals <sup>a</sup> | 3,407<br>(3,163 – 3,643) | -80<br>(-89 – -74) | -157<br>(-167 – -144) | -232<br>(-246 – -216) | -305<br>(-321 – -283) | -375<br>(-396 – -350) |
| Delayed Graft Function <sup>b</sup> | 2,941<br>(729 – 5,985) | 26<br>(-26 – 85) | 20<br>(-34 – 78) | 13<br>(-40 – 65) | 7<br>(-45 – 66) | 1<br>(-50 – 59) |
| Graft Loss <sup>b</sup> | 1,343<br>(231 – 2,852) | 47<br>(-4 – 163) | 47<br>(-3 – 163) | 47<br>(-2 – 168) | 48<br>(-3 – 170) | 49<br>(-4 – 173) |

<sup>a</sup> Rate per 10,000 kidney transplant candidates, <sup>b</sup> Rate per 10,000 deceased donor transplants

Increasing the deceased donor transplantation rate by 25% for older transplant candidates using acceptable quality kidneys results in increased quality-of-life and can be achieved in a cost-effective manner. It would cost $8,100 per QALY gained from the healthcare sector perspective (**Figure 2**). Increasing the rate of transplantation by smaller amounts costs more per QALY gained than increasing the rate by 25% because smaller increases in the rate of transplantation not only means fewer people are transplanted, but the average time to transplant is higher resulting in additional adverse outcomes and costs.

**Figure 2:**
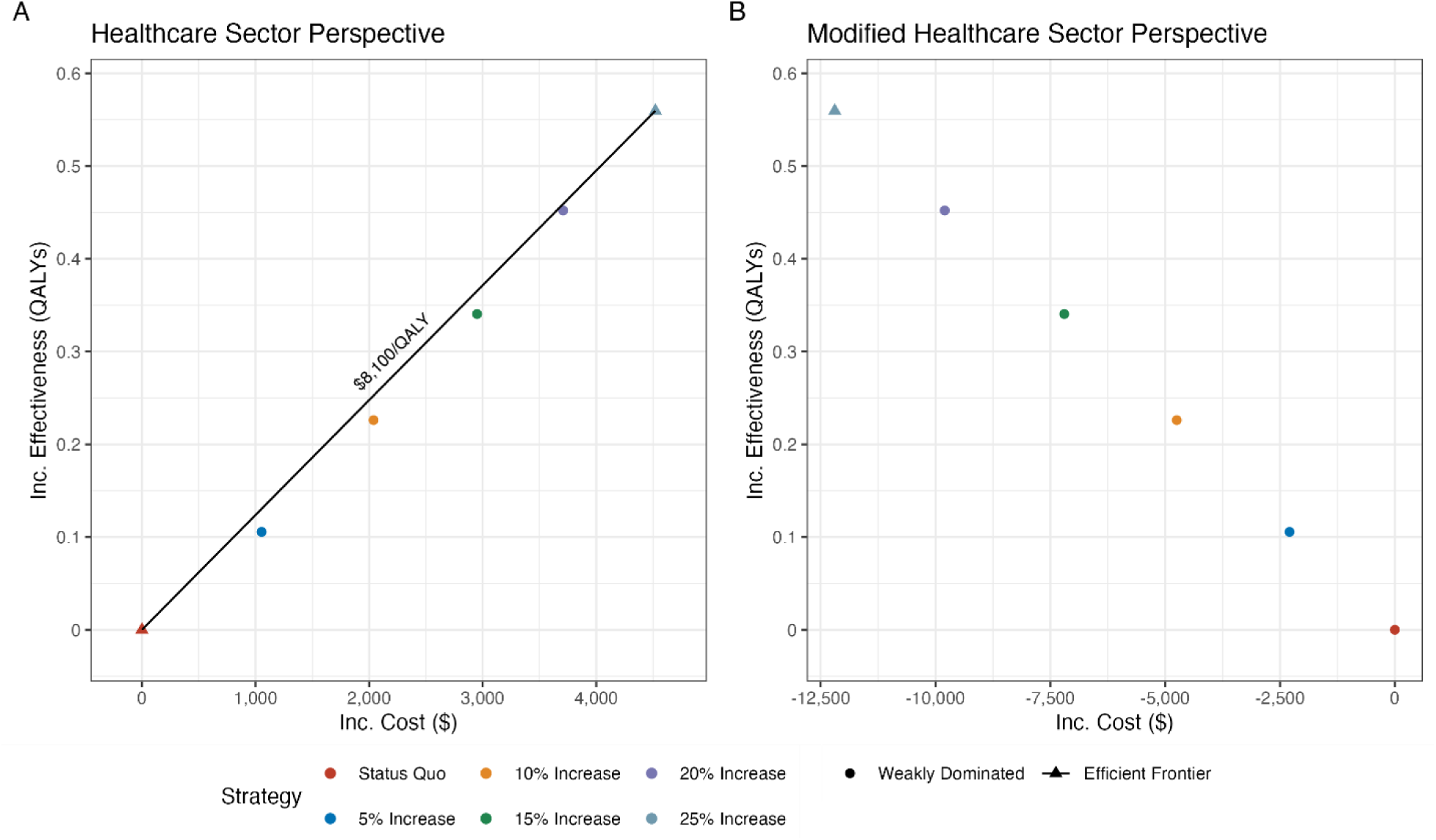
Incremental cost-effectiveness frontier for base-case results from A) healthcare sector perspective, and B) modified healthcare sector perspective. QALYs: quality-adjusted life years Inc: Incremental

From the modified healthcare sector perspective, increasing the deceased donor transplantation rate by 25% produces the most QALYs and costs less than all other options. Compared to the status quo rate of deceased donor transplantation, older candidates are expected to gain 0.56 QALYs and save $10,200.

### Subgroup Results

While older transplant candidates have shorter life expectancy and worse expected outcomes both on dialysis and post-transplant than younger candidates, our analyses show that even for individuals age ≥75 years increasing deceased donor transplant by 25% results in quality-of-life gains. It also costs $7,800 per QALY gained, well below the $100,000 per QALY gained WTP threshold from the healthcare sector perspective (**Supplemental Figure S4**).

Prior literature documents disparities in CKD, dialysis, and transplant outcomes across patients from different designated race/ethnicities.^26–33^ Notably, studies have also found that NH Black patients have better expected survival on dialysis and worse expected post-transplant outcomes compared to NH White patients.^34,35^ Increasing deceased donor transplant by 25% produces substantial health benefits for patients of all designated race/ethnicities. NH Black patients have longer than average duration on the waiting list; as such, increasing transplantation rates yields additional benefits by reducing waiting times. From the healthcare sector perspective, transplantation for all racial and ethnic subgroups have ICERs well below the $100,000 per QALY gained WTP threshold (**Supplemental Figure S5**). Gains from increasing transplantation rates would be cost-effective for all groups from the healthcare sector perspective and cost-saving from the modified healthcare sector perspective. Moreover, a strategy that reduces non-use of acceptable quality organs would enhance health equity.

Transplant candidates with diabetes have shorter life expectancy and worse health outcomes than candidates without diabetes. For individuals with diabetes, increasing the deceased donor transplantation rate by 25% resulted in quality-of-life gains, albeit by a smaller amount than those without diabetes. For those with diabetes, it would cost $9,900 per QALY gained from the healthcare sector perspective (**Supplemental Figure S6**).

### Scenario Analysis

The benefits of expanding transplantation rates by using acceptable quality deceased donor kidneys that would have otherwise gone unused involve assumptions about outcomes if these kidneys were instead transplanted. Even in scenarios analyses in which as many as 20% of transplanted kidneys are of lower quality than their KDPI implies, increasing the rate of deceased donor transplants by 25% averts 141 waitlist deaths and 375 waitlist removals. There would also be 175 additional cases of delated graft function and 103 additional graft loss events. Increasing the rate of deceased donor transplant by 25% with 20% of transplanted kidneys being of lower quality than their KDPI implies costs $8,300 per QALY gained from the healthcare sector perspective, well below the $100,000 per QALY gained WTP threshold (**Supplemental Figures S6**).

### Probabilistic Sensitivity Analysis

At a WTP threshold of $100,000 per QALY gained, increasing the rate of deceased donor transplantation by 25% was cost-effective in 100% of the PSA samples from both the healthcare sector and modified healthcare sector perspectives (**Figure 3**). It is only for WTP thresholds below $40,000 per QALY gained that the status quo (not increasing the deceased donor transplant rate) was preferred in any of the samples from the healthcare sector perspective. From the modified healthcare sector perspective, the 25% increase was cost-saving in 77% of PSA samples, even at a $0 per QALY gained threshold. We also quantified the consequences of incorrectly choosing a strategy. At a WTP threshold of $100,000 per QALY gained, choosing the status quo over increasing the rate of transplantation by 25% would result in an expected loss of $51,400 per person, for a total of $886 million, from the healthcare sector perspective, and $58,300 per person, for a total of $1 billion, from the modified healthcare sector perspective (**Supplemental Figure S13**).

**Figure 3:**
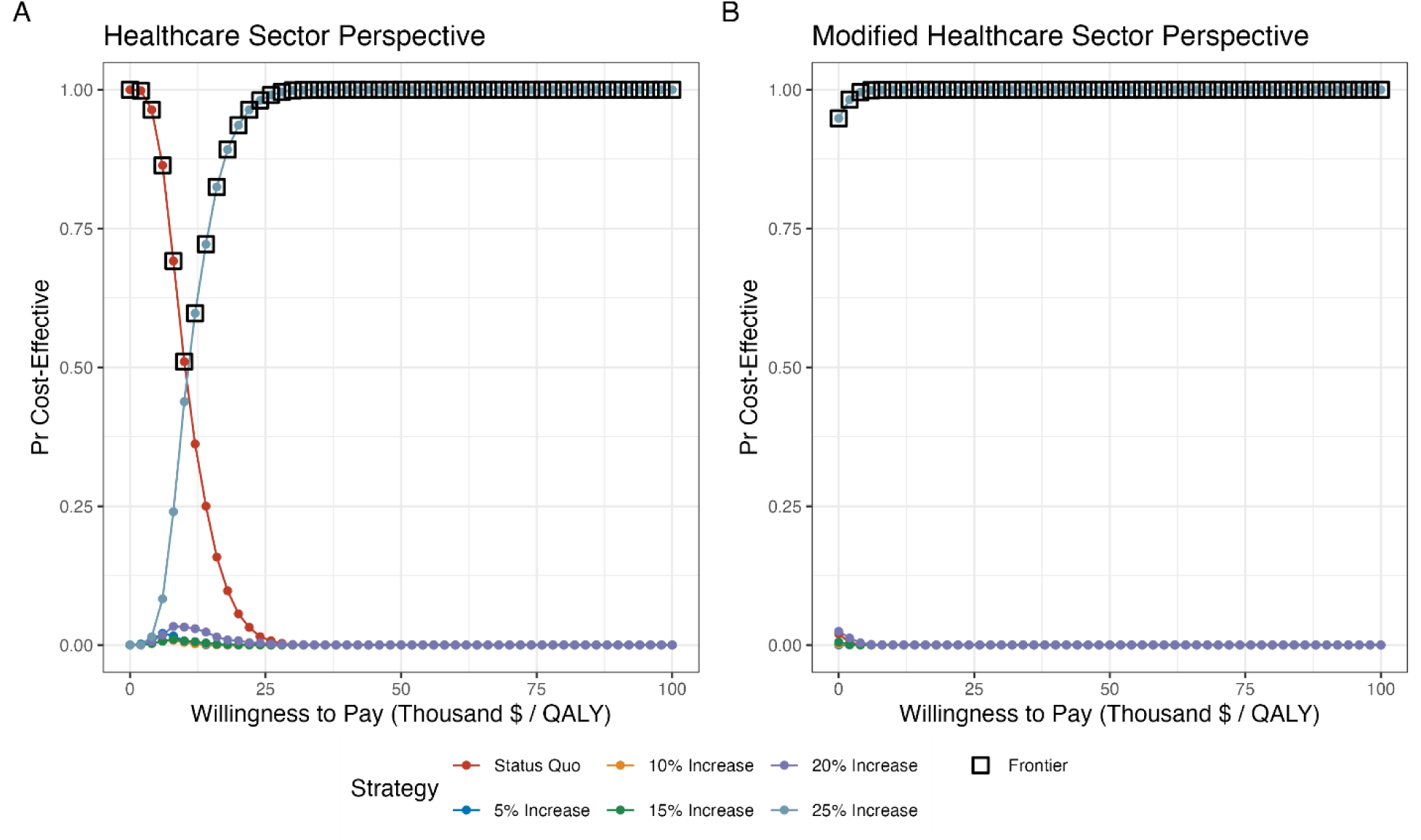
Cost-effectiveness acceptability curves and cost-effectiveness acceptability frontier from A) healthcare sector perspective, and B) modified healthcare sector perspective. QALY: quality-adjusted life year

## Discussion

Increasing the rate of deceased donor kidney transplantation for older candidates results in fewer adverse waitlist events and increases survival and health-related quality-of-life. We expect that there will be fewer waitlist deaths and waitlist removals among older candidates, with modest increases in the rate of delayed graft function and graft loss events. While current kidney allocation decisions are not made based on cost, increasing the rate of transplantation in this way can be done in a cost-effective way.

When we consider additional costs to society such as caregiver and patient time, the policy is expected to be cost saving. Using lower but acceptable quality deceased donor kidneys not only provides health benefits to the recipient but may also shorten wait times for other candidates.

Potential policies aimed at facilitating older candidates’ access to more available and acceptable quality kidneys should be considered. Our analysis does not address the issues that ESKD patients face related to financial and practical barriers in access to care. It also does not suggest that more older candidates should be listed for transplantation. The current system is constrained in such a way that patient preferences are unlikely to be honored. While many older patients would prefer a kidney transplant if available, the decision might be informed by a variety of factors, including how well they are faring on dialysis, underlying diseases that might modify the risk of complications of immunosuppression, or other factors unique to each individual patient (e.g., options for home dialysis, availability of sites for hemodialysis vascular access).^36^ Older transplant candidates have shown a willingness to accept lower kidney quality in order to have less wait time, hence the reason our analysis focuses on this population.^8^ However, we believe that access to any acceptable quality kidneys should be extended to any candidate who expresses this preference. If patients are informed of the risks associated with receiving a lower quality donor kidney, transplant centers should not be penalized for honoring their preferences. The most recent meeting on stakeholder perspectives resulted in recommendations that include shared decision making around whether to accept offers for medically complex donors.^37^ We believe that there is potential to create a policy that allows older candidates access to such organs without being subject to performance metrics. One such policy involves exempting older candidates who are willing to accept offers for lower quality donor kidneys from transplant center performance metrics. Exempting these cases would reduce the incentives for transplant centers to accept only the highest quality donor kidneys (and approve only the healthiest of patients) to optimize performance metrics. Given that performance metrics are currently under review, there is an opportunity to tailor performance metrics that allow patient preferences to be honored.

A strength of our study lies in the ability to use patient level data that is representative of older waitlisted kidney transplant candidates. People’s risks are determined by a complex combination of factors like their time on the waitlist, age at transplant, comorbidities, and quality of the kidney they receive. By using individual-level data on all older transplant candidates in the United States, the model can appropriately reflect the heterogeneous risks of outcomes for individuals. We also perform a full probabilistic analysis based on established calibration methods.^10,38^ This allows us to reflect the uncertainty in predicted outcomes consistent with the empirical data. Notably, even with substantial uncertainty and the inclusion of frequently omitted substantial costs, our findings about the use of lower quality kidneys being cost-effective or cost-saving are robust. Unlike earlier cost-effectiveness studies of different policies in kidney transplantation, we included a more comprehensive assessment of costs. These costs include those related to organ acquisition and opportunity costs experienced by patients and caregivers.

We elected not to consider downstream effects of this policy on younger transplant candidates for simplicity’s sake. If we were to consider effects on younger transplant candidates, we would expect to see downstream benefits, since placing more donor kidneys in the pool would lead to shorter waiting times for younger candidates relative to the status quo. Risks for simulated individuals in our model depend on race/ethnicity but do not depend on other socioeconomic measures. The reason for this is the underlying waitlist and transplant data do not have sufficient information on socioeconomic measures for us to include in the estimated risk equations we used. As socioeconomic factors can reduce access to and bias experienced in healthcare settings, our model fails to capture these effects. To the extent that designated race/ethnicity correlates with these socioeconomic factors, modeled differences in outcomes across race/ethnicity may capture some variability related to socioeconomic factors, as well as other social determinants of health, including system racism and other forms of discrimination.

Allowing older kidney transplant candidates access to potentially usable, lower but acceptable quality deceased donor kidneys that are currently going unused would improve health outcomes in a cost-effective manner. Doing so would also increase health equity, as currently, older transplant candidates receive a disproportionally low number of transplants, leading to worse health outcomes, despite stated preferences for accepting lower quality kidneys to shorten these waits. Our analysis suggests that decision makers updating transplant center performance metrics should consider policies that could use currently underutilized kidneys to increase transplantation in older patients and any other patients with similar preferences.

## Data Availability

All data produced in the present study are available upon request to the authors. Model code can be found on our GitHub page.

https://github.com/mbkauf/OlderKidneyTransplantSim

## Acknowledgments

We would like to acknowledge the Stanford Transplant Clinical Research and the Stanford Data Science × Decision Science research hub for providing insightful commentary on the work. Some of the computing for this project was performed on the Sherlock cluster. We would like to thank Stanford University and the Stanford Research Computing Center for providing computational resources and support that contributed to these research results.

## Role of Funding Source

The funders did not have any role in the study design, analysis, or preparation of the manuscript.

## Notes

### Competing Interest Statement

GMC reports royalties or licenses from Elsevier, Brenner and Rectors The Kidney; Consulting fees for Trial Steering Committees from Akebia, AstraZeneca, Sanifit, and Vertex; Consulting fees for advisory board from Alebund, Ardelyx, CalciMedica, Miromatrix, Panoramic, Renibus, Toku, and Unicycive; Support for attending meetings and/or travel from AstraZeneca, CSL Behring, and Vertex; Participation on a Data Safety Monitoring Board or Advisory Board from Aethlon, Bayer, Mineralys, and ReCor; Leadership or fiduciary role in other board, society, committee or advocacy group (unpaid) with Satellite Healthcare; Stock options from CloudCath, Eliaz Therapeutics, Outset, Renibus, Unicycive. Other authors report no conflicts of interest.

### Funding Statement

This project was supported by grant T32HS026128 from the Agency for Healthcare Research and Quality. The content is solely the responsibility of the authors and does not necessarily represent the official views of the Agency for Healthcare Research and Quality.

