## Supplementary Materials for "Cost-effectiveness of transplanting older candidates with acceptable quality deceased donor kidneys"

### Table of Contents

Methods S1: Cost Parameters and Calculations ........ **2-3**

Methods S2: Health-Related Quality of Life Calculations ........ **3-5**

Methods S3: Model Calibration ........ **5-8**

Methods S4: Impact inventory and CHEERS checklist ........ **8-13**

Methods S5: Analysis Methodologies ........ **13**

Appendix Results ........ **13-20**

Appendix Citations ........ **23**

#### Methods S1: Cost Parameters and Calculations

The United States Renal Data System (USRDS) Annual Report provides total costs for dialysis, transplantation, and post-transplant costs which also include complications (including DGF and graft failure) that the model explicitly accounts for. These are based on Medicare costs, including Part D.[1] To avoid double counting the cost of complications, we disaggregated the total cost of transplantation into the initial hospitalization costs without complications and the cost of DGF. The estimated difference between hospital costs for recipients with and without DGF from Almond et al., 1991, is $15,760.[2] This value is 1989 USD, which we adjust to 2022 USD using the PCE-Health total, is $41,592.[3] Then we estimate the cost of deceased donor transplantation without delayed graft function (DGF). We start with our estimate cost of deceased donor transplantation from Axelrod et al., 2017.[4] The estimate provided, $106,675, is in 2013 USD. We then adjust this estimate to 2022 USD using the PCE-Health total over that period, which comes out to $125,127.[3] Next, we multiply the fraction of deceased donor kidney transplant recipients who experienced DGF by our DGF cost estimate.[2] Our final cost estimate for deceased donor transplantation is $125,127 – (0.251 * $41,592) = $114,688. We derived the cost of graft failure from the post-transplantation costs for a year in which graft failure occurs, by assuming that, on average, in the year of graft failure, the recipient has a functioning graft for 6 months, has the acute graft loss event during the next month, and 5 subsequent months of dialysis.[5]

For our probabilistic sensitivity analysis, the costs for each parameter set were drawn from the distributions described in **Table S1**. For costs that are drawn from gamma distributions, we use Nelder-Mead optimization to calibrate the rate and shape parameters. We minimize the sum of squared errors of the mean and standard deviation of samples from a gamma distribution and our mean and standard deviation targets. For costs that are drawn from a uniform distribution, we use +/- 20% of the point estimates.

Many of the cost inputs did not report uncertain ranges, standard deviations, or standard errors. Our approach to determining the standard deviations that we use as targets in the optimization algorithm is dependent on the source. For dialysis and post-transplant costs, we used the standard deviation of all the age-specific costs as the standard deviation for each individual age-specific cost. We use the same approach for the one-time graft loss event costs.

For our transplant costs we used the HCUPnet database as a means of determining the variation.[6] The standard error of DRG code 652, Kidney Transplant" was 3.03% of the point estimate. We assume that the standard deviation of both deceased donor and living donor transplants to be the same percentage of our point estimates. Because our DGF costs are a difference in means, we use the formula for the standard deviations of the difference of sample means:

$$\sigma_d = \sqrt{\sigma_1^2/n_1 + \sigma_2^2/n_2}$$

For caregiver time for patients on dialysis we fix the wage estimates and use the standard deviation of the caregiving hours. We multiply the standard deviation of the hours per month of caregiving time by the BLS estimate.

**Table S1** shows the cost parameters along with the distributions that are sampled from for the probabilistic sensitivity analysis.

Table S1: Cost Inputs

| Cost | Value | Distribution | Source(s) |
|---|---|---|---|
| **Healthcare Sector Perspective** | | | |
| Dialysis w/ Part D 65-69 (monthly) | $8,791 | Gamma(shape = 3312.222, rate = 0.377) | 2021 USRDS Annual Data Report |
| Dialysis w/ Part D 70-74 (monthly) | $8,675 | Gamma(shape = 3206.55, rate = 0.37) | 2021 USRDS Annual Data Report |
| Dialysis w/ Part D 75-79 (monthly) | $8,553 | Gamma(shape = 3999.94, rate = 0.468) | 2021 USRDS Annual Data Report |
| Dialysis w/ Part D 80-84 (monthly) | $8,672 | Gamma(shape = 3211.89, rate = 0.37) | 2021 USRDS Annual Data Report |
| Dialysis w/ Part D 85+ (monthly) | $8,387 | Gamma(shape = 2993.366, rate = 0.357) | 2021 USRDS Annual Data Report |
| Post-Transplant w/ Part D 65-69 (monthly) | $3,520 | Gamma(shape = 476.943, rate = 0.135) | 2021 USRDS Annual Data Report |
| Post-Transplant w/ Part D 70-74 (monthly) | $3,498 | Gamma(shape = 470.804, rate = 0.135) | 2021 USRDS Annual Data Report |

| | | | |
|---|---|---|---|
| Post-Transplant w/ Part D 75-79 (monthly) | $3,604 | Gamma(shape = 1206.225, rate = 0.335) | 2021 USRDS Annual Data Report |
| Post-Transplant w/ Part D 80-84 (monthly) | $3,419 | Gamma(shape = 448.123, rate = 0.131) | 2021 USRDS Annual Data Report |
| Post-Transplant w/ Part D 85+ (monthly) | $3,182 | Gamma(shape = 1509.844, rate = 0.474) | 2021 USRDS Annual Data Report |
| Deceased Donor Transplantation (one-time) | $114,687 | Gamma(shape = 57402.46, rate = 0.5005) | Axelrod et al., 2017 |
| Organ Acquisition Cost Center (one-time) | $112,318 | Gamma(shape = 11.232, rate = 0.0001) | Cheng et al., 2022 |
| Delayed Graft Function (one-time) | $41,592 | Gamma(shape = 24.925, rate = 0.0006) | Almond et al., 1991 |
| Graft Failure (one-time) | $85,989 | Gamma(shape = 8.599, rate = 0.0001) | 2021 USRDS Annual Data Report |
| Living Donor Transplant (one-time) | $111,976 | Gamma(shape = 56062.28, rate = 0.5007) | Axelrod et al., 2017 |
| Post-Waitlist Removal (multiplier) | 1.2 | Uniform(1.0, 1.4) | Assumed |
| **Societal Perspective** | | | |
| Caregiver Time | | | |
| Dialysis (monthly) | $5,867 | Gamma(shape = 2934.9, rate = 0.05) | Liu et al., 2022, BLS May 2021 |
| Post-Transplant (monthly) | $1,141 | Gamma(shape = 2.0905, rate = 0.0018) | Langa et al., 2004, BLS May 2021 |
| Patient Time | | | |
| Dialysis (monthly) | $1,611 | Uniform(1289, 1933) | BLS May 2021, Expert Opinion |
| Post-Transplant: Month 1 (monthly) | $372 | Uniform(298, 446) | BLS May 2021, Expert Opinion |
| Post-Transplant: Month 2-3 (monthly) | $186 | Uniform(149, 223) | BLS May 2021, Expert Opinion |
| Post-Transplant: Month 4+ (monthly) | $62 | Uniform(50, 74) | BLS May 2021, Expert Opinion |

Note: Post-Transplant Removal multiplier is applied to dialysis costs to reflect that those removed from the waitlist are likely to have increased healthcare needs.

#### Methods S2: Health-Related Quality of Life Calculations

The health-related quality of life weight calculations we are derived using two sources, Hanmer et al., 2006 and Wyld et al., 2012.[7,8] Wyld et al., 2012 is a meta-analysis of studies that estimate quality of life weights for kidney transplant candidates and recipients. From this paper, we use Table 4, which presents the quality-of-life weights from longitudinal studies that estimate the weights pre- and post-transplant (**Table S2**).

##### Table S2. Longitudinal health-related quality-of-life studies of transplant recipients

| Study | Utility Elicitation Instrument | Number of Patients | Utility | | | | | Mean Age |
|---|---|---|---|---|---|---|---|---|
| | | | Pre-Transplant | Post-Transplant | | | | |
| | | | | 0—3 mo | 4—8 mo | 9-12 mo | 13-24 mo | |
| Balaska et al. (20] | SF-36 | 85 | 0.35 | | | 0.6 | | 43.8 |
| Laupacis et al. (21] | TTO | 131 | 0.57 | 0.71 | 0.75 | 0.74 | 0.7 | 42 |
| Oberbauer et al. (22), group 1 | SF-36 | 183 | | 0.61 | | 0.62 | 0.62 | 43.9 |
| Oberbauer et al. (22), group 2 | SF-36 | 178 | | 0.61 | | 0.6 | 0.6 | 45.2 |
| Painter et al. (23), group 1 | SF-36 | 14 | | 0.59 | | 0.58 | | 48.3 |
| Painter et al. (23), group 2 | SF-36 | 9 | | 0.67 | | 0.69 | | 46.8 |

| | | | | | | | | |
|---|---|---|---|---|---|---|---|---|
| Perez San Gregorio et al. (24) | SF-36 | 28 | 0.59 | 0.57 | 0.63 | 0.64 | | 40.61 |
| Pinson et al. (25) | SF-36 | 24 | 0.58 | 0.56 | | | | 44 |
| Ravagnani et al. [26) | SF-36 | 17 | 0.57 | | | | 0.61 | 37.9 |
| Rodrigue et al. (27) | SF-36 | 31 | 0.56 | 0.57 | 0.62 | 0.65 | | |
| Russell et al. [28] | TTO | 27 | 0.41 | | | 0.74 | | 41.9 |
| Weighted Average | | 727 | 0.50 | 0.63 | 0.71 | 0.64 | 0.63 | 43.64 |

Adapted from Wyld et al., 2012 Table 4.

We derived these weights from studies with an average age younger than our population of interest. Because younger populations on average have higher health-related quality of life, we scaled these weights downward for our older population given that quality of life is lower in older ages.[9] We first calculated the difference between the quality of life weights of the population of transplant candidates and recipients and the general population of the same age. We then applied those differences to the age- and sex-specific quality of life weights that aligned with our population of candidates 65 and older.[9] To estimate age-, sex-, and time-since-transplant-specific weights, we used the Hanmer et al., 2006 EQ-5D estimates.[7] The studies for transplant related weights have an average age of about 44. We take the difference between these estimates and those of the general US population from ages 40-49 for males and females. Using the time-since-transplant differences from the general population, we subtract that difference from the age ranges relevant to our study population (60-69, 70-79, and 80-89). These are our age-, sex-, and time-since-transplant-specific weights. We assume that those 90 and older have the same weights as those 80-89.

For our probabilistic sensitivity analysis, we draw from a beta distribution for our health-related quality of life weights. We estimate the beta distribution parameters using the estimated weights and the total sample size of the studies. They are calculated as:

$$\alpha = nw$$
$$\beta = n(1 - w)$$

where $n$ is the sample size and $w$ is the quality-of-life weight. Using the sample size of the studies in **Table S2** ($n = 727$), the $\alpha$ and $\beta$ parameters are shown in **Table S3.**

#### Table S3 Health-related Quality of Life Weight Distribution Parameters

| | Male | | | | | Female | | | | |
|---|---|---|---|---|---|---|---|---|---|---|
| Age | Pre-Tx | 0—3 mo | 4—8 mo | 9-12 mo | 13-24 mo | Pre-Tx | 0—3 mo | 4—8 mo | 9-12 mo | 13-24 mo |
| **Alpha** | | | | | | | | | | |
| 60-69 | 334 | 422 | 480 | 436 | 429 | 327 | 414 | 480 | 429 | 422 |
| 70-79 | 305 | 393 | 458 | 407 | 400 | 298 | 385 | 451 | 400 | 393 |
| 80+ | 291 | 378 | 443 | 393 | 385 | 269 | 356 | 414 | 364 | 356 |
| **Beta** | | | | | | | | | | |
| 60-69 | 395 | 306 | 244 | 294 | 301 | 398 | 309 | 248 | 298 | 304 |
| 70-79 | 422 | 333 | 272 | 322 | 328 | 427 | 338 | 277 | 327 | 334 |
| 80+ | 437 | 348 | 286 | 336 | 343 | 461 | 373 | 311 | 361 | 368 |

#### Table S4: Health-related Quality of Life Inputs

| | Age | | | | | |
|---|---|---|---|---|---|---|
| **QALY** | **All** | **65-69** | **70-79** | **80+** | **Distribution** | **Source(s)** |
| Dialysis | | | | | | |

| | | | | | | |
|---|---|---|---|---|---|---|
| Male | | 0.46 | 0.42 | 0.40 | Beta(alpha = (334, 305, 291) beta = (393, 422, 436)) | Hanmer et al., 2006, Wyld et al., 2012 |
| Female | | 0.43 | 0.39 | 0.34 | Beta(alpha = (313, 284, 247) beta = 414, 443, 480)) | Hanmer et al., 2006, Wyld et al., 2012 |
| Post-Transplant (0-3 Months) | | | | | | |
| Male | | 0.58 | 0.54 | 0.52 | Beta(alpha = (422, 393, 378) beta = (305, 334, 349)) | Hanmer et al., 2006, Wyld et al., 2012 |
| Female | | 0.55 | 0.51 | 0.46 | Beta(alpha = (400, 371, 334) beta = (327, 356, 393)) | Hanmer et al., 2006, Wyld et al., 2012 |
| Post-Transplant (4-8 Months) | | | | | | |
| Male | | 0.66 | 0.63 | 0.61 | Beta(alpha = (480, 458, 443) beta = (247, 269, 284)) | Hanmer et al., 2006, Wyld et al., 2012 |
| Female | | 0.64 | 0.60 | 0.55 | Beta(alpha = (465, 436, 400) beta = (262, 291, 327)) | Hanmer et al., 2006, Wyld et al., 2012 |
| Post-Transplant (9-12 Months) | | | | | | |
| Male | | 0.60 | 0.56 | 0.54 | Beta(alpha = (436, 407, 393) beta = (291, 320, 334)) | Hanmer et al., 2006, Wyld et al., 2012 |
| Female | | 0.57 | 0.53 | 0.48 | Beta(alpha = (414, 385, 393) beta = (313, 342, 378)) | Hanmer et al., 2006, Wyld et al., 2012 |
| Post-Transplant (13+ Months) | | | | | | |
| Male | | 0.59 | 0.55 | 0.53 | Beta(alpha = (429, 400, 385) beta = (298, 327, 342)) | Hanmer et al., 2006, Wyld et al., 2012 |
| Female | | 0.56 | 0.52 | 0.47 | Beta(alpha = (407, 378, 342) beta = (320, 349, 385)) | Hanmer et al., 2006, Wyld et al., 2012 |
| Delayed Graft Function (one-time) | 0 | | | | | Assumed |
| Graft Failure (one-time) | 0 | | | | | Assumed |

Note: we assume that patients who experience delayed graft function and graft failure derive 0 QALYs for the month in which these events occur.

#### Methods S3: Model Calibration

After the publication of the manuscript that describes our model development and calibration, we sought to improve the fit of the model with a focus on post-transplant outcomes.[10] We changed our goodness-of-fit metric from a sum of squared errors to a likelihood-based approach. The targets are derived from the data that was held out from equation development. For the survival equations, we tested two methods of calibration. The first was to use points along the Kaplan-Meier curves, assuming a normal distribution around the point estimates. The second method was to fit parametric regressions without covariates to the held-out data and use the auxiliary parameters as the targets. We then assumed a multivariate normal distribution around the auxiliary parameters. For the 30-day outcomes that are estimated using a multinomial logistic regression, our targets assumed a Dirichlet distribution. For death the same day as graft loss, which we estimate using a logistic regression, we assumed a binomial distribution. We also added additional calibration targets for the patient characteristics at the time of transplant. These characteristics include age at transplant, sex, race/ethnicity, blood type, years on dialysis before transplant, diabetes history, chronic obstructive pulmonary disease (COPD) history, peripheral vascular disease (PVD) history, angina/ coronary artery disease (CAD) history, peak calculated panel reactive antibodies (cPRA), and Organ Procurement and Transplantation Network (OPTN) region. We assumed normal distributions for each of these characteristics.

Another change to our calibration method was to conduct the calibration in two stages: waitlist outcomes and post-transplant outcomes. Once we have a sufficient number of parameter sets that fit the waitlist outcomes well, we sample from those parameter sets of waitlist equations and combine them with parameter sets for the post-transplant outcome equations. We do this because the accuracy of the post-transplant outcomes is dependent on first predicting the correct mix of patients who receive a deceased donor kidney. We generated 100,000 parameter sets and identified 64 parameter sets that we deemed to be acceptable.

**Table S5** shows patient characteristics at the time of transplant for the observed data compared to our calibrated parameter sets. **Figures S1-S3** show how the simulated outcomes compare to those in the held-out observed data. We can see that we have good fit to the observed data. Compared to the original calibrated parameter sets, we see the most improvement for our death after graft loss and 30-day outcomes equations.

Table S5: Patient characteristics at the time of transplant

| | Observed | Simulated (95% CrI) |
|---|---|---|
| Age at Transplant | 69.3 | 70.44 (70.33 - 70.56) |
| Sex | 0.62 | 0.62 (0.61 - 0.62) |
| White | 0.51 | 0.49 (0.48 - 0.5) |
| Black | 0.25 | 0.28 (0.27 - 0.28) |
| Hispanic | 0.14 | 0.13 (0.13 - 0.13) |
| Other Race/Ethnicity | 0.1 | 0.1 (0.1 - 0.11) |
| Blood A | 0.37 | 0.36 (0.35 - 0.36) |
| Blood AB | 0.05 | 0.05 (0.05 - 0.05) |
| Blood B | 0.14 | 0.14 (0.13 - 0.15) |
| Blood O | 0.44 | 0.45 (0.44 - 0.46) |
| Years on Dialysis Before Transplant | 3.97 | 3.25 (3.19 - 3.32) |
| History of Diabetes | 0.51 | 0.51 (0.5 - 0.52) |
| History of COPD | 0.01 | 0.01 (0.01 - 0.01) |
| History of PVD | 0.12 | 0.12 (0.12 - 0.12) |
| History of Angina/CAD | 0.55 | 0.59 (0.58 - 0.6) |
| cPRA | 0.17 | 0.17 (0.17 - 0.17) |
| OPTN 1 | 0.04 | 0.04 (0.04 - 0.04) |
| OPTN 2 | 0.14 | 0.13 (0.12 - 0.15) |
| OPTN 3 | 0.14 | 0.14 (0.14 - 0.14) |
| OPTN 4 | 0.07 | 0.06 (0.06 - 0.07) |
| OPTN 5 | 0.17 | 0.16 (0.15 - 0.17) |
| OPTN 6 | 0.05 | 0.06 (0.06 - 0.06) |
| OPTN 7 | 0.07 | 0.06 (0.05 - 0.06) |
| OPTN 8 | 0.07 | 0.08 (0.08 - 0.09) |
| OPTN 9 | 0.07 | 0.07 (0.06 - 0.07) |
| OPTN 10 | 0.08 | 0.08 (0.08 - 0.09) |
| OPTN 11 | 0.1 | 0.12 (0.11 - 0.12) |

We compared the observed versus simulated patient characteristics and the 95% credible interval.

#### Figure S1: Model calibration Kaplan-Meier curves for waitlist outcomes.

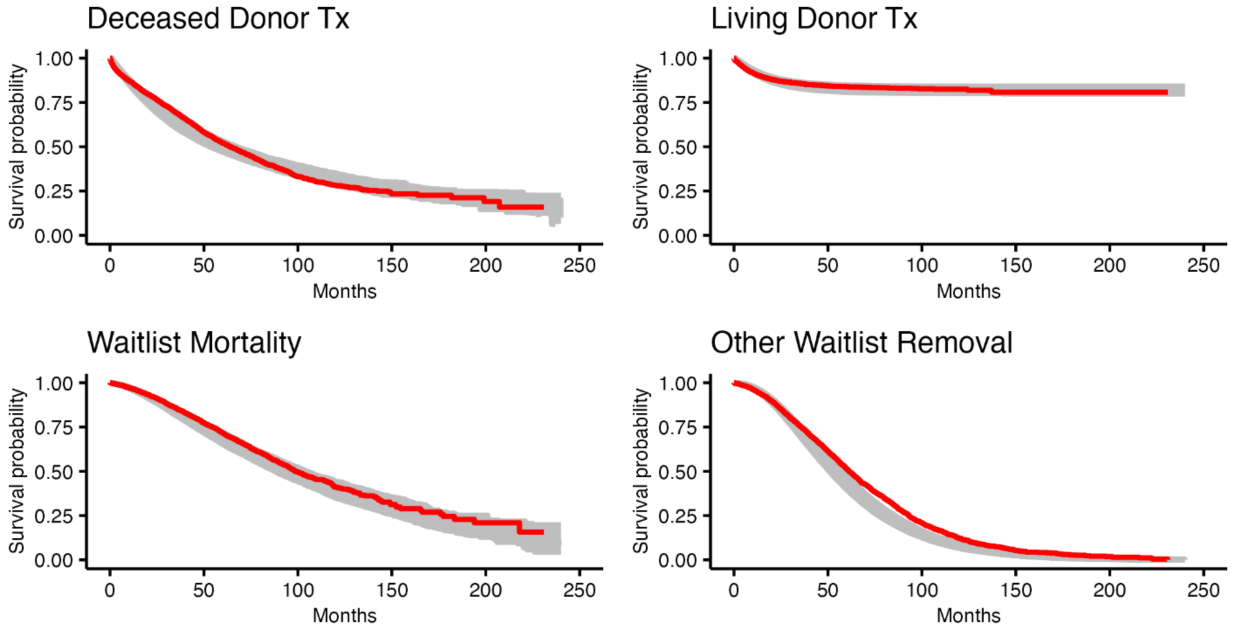


The Kaplan-Meier curves show simulated versus observed survival. The gray curves each represent 1 of the simulated best-fitting parameter sets, and the red curve represents the observed data.

#### Figure S2: Model calibration Kaplan-Meier curves for post-transplant outcomes

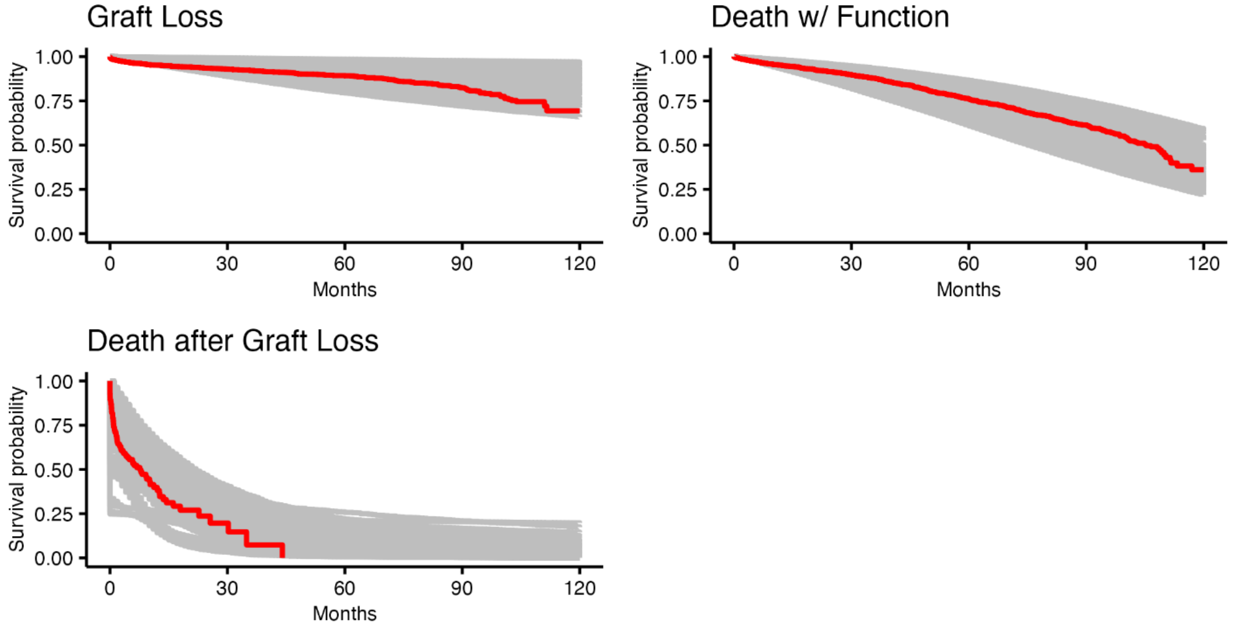


The Kaplan-Meier curves show simulated versus observed survival. The gray curves each represent 1 of the simulated best-fitting parameter sets, and the red curve represents the observed data.

#### Figure S3: Model calibration plots for 30-day outcomes and death at graft loss

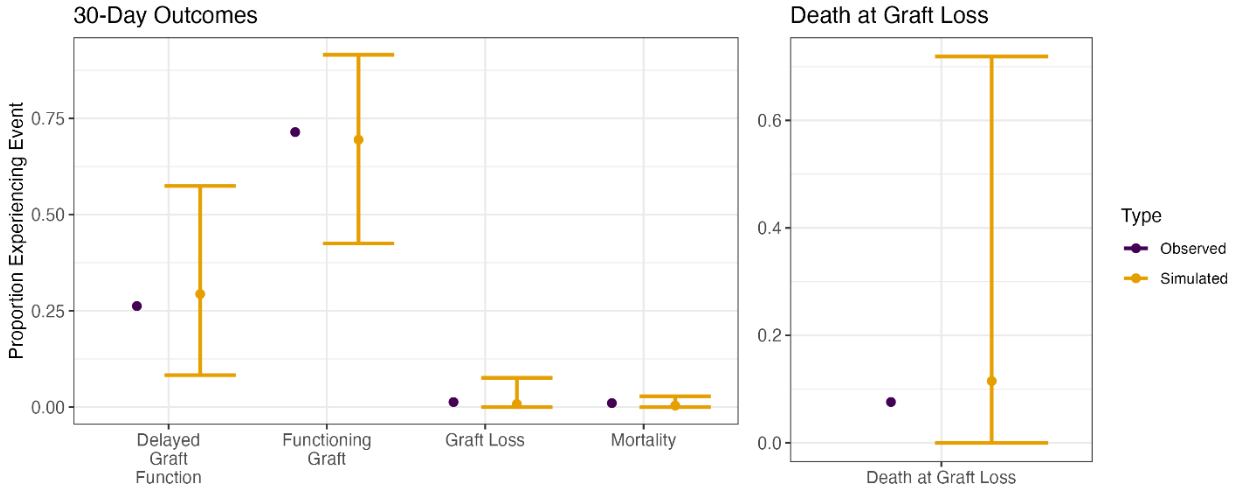


The 30-day outcomes were predicted using a multinomial logistic regression while death at graft loss was predicted using logistic regression. The purple represents the calibration targets while yellow represents the simulated mean and 95% credible intervals from the calibrated parameter sets.

#### Methods S4: Impact inventory and CHEERS checklist

##### Table S6: Impact Inventory

| Sector | Type of Impact | Included in this Reference Case Analysis From…Perspective? | | Notes on Sources of Evidence |
|---|---|---|---|---|
| | | Health Care Sector | Societal | |
| Formal Health Care Sector | | | | |
| Health | Health Outcomes (effects) | | | |
| | Longevity effects | Y | Y | Model equations account for life expectancy. Life expectancy after exiting model comes from 2021 USRDS annual data report |
| | Health-related QoL effects | Y | Y | QALY estimates from Wyld et al. 2012 for transplant vs dialysis. QALY estimates are adjusted for age and sex |
| | Other health effects (eg, adverse events) | Y | Y | Model equations track number of events for: transplants, waitlist and post-transplant mortality, waitlist removals, delayed graft function, and graft loss |

| | | | | |
|---|---|---|---|---|
| | Medical Costs | | | |
| | Paid for by third-party payers | Y | Y | Medicare spending by modality and event type from 2021 USRDS annual data report |
| | Paid for by patients out-of-pocket | N | N | Only using Medicare spending as for cost estimates |
| | Future related medical costs (payers and patients) | Y | Y | 2021 USRDS Annual Data Report has related medical costs |
| | Future unrelated medical costs (payers and patients) | Y | Y | 2021 USRDS Annual Data Report has related medical costs |
| Informal Health Care Sector | | | | |
| Health | Patient-time costs | NA | Y | Dialysis 3 Times a week, 4 hours each Post-transplant time based on schedule of office visits and lab draws. |
| | Unpaid caregiver-time costs | NA | Y | Substantial caregiver-time pre-transplant, lower costs post-transplant |
| | Transportation costs | NA | N | Included in caregiver-time costs |
| Non-Health Care Sectors | | | | |
| Productivity | Labor market earnings lost | NA | N | Older population assumed to be of retirement age |
| | Cost of unpaid lost productivity due to illness | NA | N | Older population assumed to be of retirement age |
| | Cost of uncompensated household production | NA | N | Older population assumed to be of retirement age |
| Consumption | Future consumption unrelated to health | NA | N | Not considered in our study |
| Social Services | Cost of social services as a part of intervention | NA | N | Not applicable |
| Legal or Criminal Justice | Number of crimes related to intervention | NA | N | Not applicable |
| | Cost of crimes related to intervention | NA | N | Not applicable |
| Education | Impact of intervention on educational achievement of population | NA | N | Not applicable to the older population |
| Housing | Cost of intervention on home improvements | NA | N | Assume to be in-center dialysis |
| Environment | Production of toxic waste pollution by intervention | NA | N | Not applicable |

| | | | | |
|---|---|---|---|---|
| Other (specify) | Other impacts | NA | N | No other impacts identified |

#### Table S7: CHEERS Checklist

| Topic | No. | Item | Location where item is reported |
|---|---|---|---|
| **Title** | | | |
| Title | 1 | Identify the study as an economic evaluation and specify the interventions being compared. | Title, Page 1 |
| **Abstract** | | | |
| Abstract | 2 | Provide a structured summary that highlights context, key methods, results, and alternative analyses. | Abstract, Pages 1 |
| **Introduction** | | | |
| **Background and objectives** | 3 | Give the context for the study, the study question, and its practical relevance for decision making in policy or practice. | Introduction, Pages 2-3 |
| **Methods** | | | |
| **Health economic analysis plan** | 4 | Indicate whether a health economic analysis plan was developed and where available. | Not reported |
| **Study population** | 5 | Describe characteristics of the study population (such as age range, demographics, socioeconomic, or clinical characteristics). | Methods, Model Description Subsection |
| **Setting and location** | 6 | Provide relevant contextual information that may influence findings. | Methods, Model Description Subsection |
| **Comparators** | 7 | Describe the interventions or strategies being compared and why chosen. | Methods, Interventions Subsection |
| **Perspective** | 8 | State the perspective(s) adopted by the study and why chosen. | Methods, Analysis Subsection |
| **Time horizon** | 9 | State the time horizon for the study and why appropriate. | Methods, Analysis Subsection |
| **Discount rate** | 10 | Report the discount rate(s) and reason chosen. | Methods, Analysis Subsection |
| **Selection of outcomes** | 11 | Describe what outcomes were used as the measure(s) of benefit(s) and harm(s). | Methods, Analysis Subsection |
| **Measurement of outcomes** | 12 | Describe how outcomes used to capture benefit(s) and harm(s) were measured. | Methods, Analysis Subsection |

| Topic | No. | Item | Location where item is reported |
|---|---|---|---|
| **Valuation of outcomes** | 13 | Describe the population and methods used to measure and value outcomes. | Methods, Data and Sources Subsection |
| **Measurement and valuation of resources and costs** | 14 | Describe how costs were valued. | Methods, Data and Sources Subsection, Costs Subsubsection |
| **Currency, price date, and conversion** | 15 | Report the dates of the estimated resource quantities and unit costs, plus the currency and year of conversion. | Methods, Analysis Subsection |
| **Rationale and description of model** | 16 | If modelling is used, describe in detail and why used. Report if the model is publicly available and where it can be accessed. | Methods, Model Description Subsection |
| **Analytics and assumptions** | 17 | Describe any methods for analyzing or statistically transforming data, any extrapolation methods, and approaches for validating any model used. | Methods and Supplemental Materials |
| **Characterizing heterogeneity** | 18 | Describe any methods used for estimating how the results of the study vary for subgroups. | Methods, Analysis Subsection |
| **Characterizing distributional effects** | 19 | Describe how impacts are distributed across different individuals or adjustments made to reflect priority populations. | Not reported |
| **Characterizing uncertainty** | 20 | Describe methods to characterize any sources of uncertainty in the analysis. | Methods, Analysis subsection |
| **Approach to engagement with patients and others affected by the study** | 21 | Describe any approaches to engage patients or service recipients, the general public, communities, or stakeholders (such as clinicians or payers) in the design of the study. | Not reported |
| **Results** | | | |
| **Study parameters** | 22 | Report all analytic inputs (such as values, ranges, references) including uncertainty or distributional assumptions. | Methods, Data and Sources subsection, and Appendix |
| **Summary of main results** | 23 | Report the mean values for the main categories of costs and outcomes of interest and summarize them in the most appropriate overall measure. | Results, Base case results subsection |

| Topic | No. | Item | Location where item is reported |
| --- | --- | --- | --- |
| **Effect of uncertainty** | 24 | Describe how uncertainty about analytic judgments, inputs, or projections affect findings. Report the effect of choice of discount rate and time horizon, if applicable. | Results, Scenario Analysis subsection and Probabilistic Sensitivity Analysis subsection |
| **Effect of engagement with patients and others affected by the study** | 25 | Report on any difference patient/service recipient, general public, community, or stakeholder involvement made to the approach or findings of the study | Not reported |
| **Discussion** | | | |
| **Study findings, limitations, generalizability, and current knowledge** | 26 | Report key findings, limitations, ethical or equity considerations not captured, and how these could affect patients, policy, or practice. | Discussion |
| **Other relevant information** | | | |
| **Source of funding** | 27 | Describe how the study was funded and any role of the funder in the identification, design, conduct, and reporting of the analysis | Methods, Role of Funding Source subsection |
| **Conflicts of interest** | 28 | Report authors conflicts of interest according to journal or International Committee of Medical Journal Editors requirements. | Conflicts of interest section |

#### Methods S5: Analysis Methodology

In our base case and scenario analyses, we simulated a population of 10,000,000 individuals per natural history parameter set from calibration to minimize stochastic noise due to simulating rare events. We calculated the difference in cost and quality of life weights for each parameter set, and then averaged over all the sets to determine cost-effectiveness and ICERs.[11]

We explored model uncertainty by running a probabilistic sensitivity analysis (PSA). For each of the 64 parameter sets from calibration, we combined them with 100 samples from the uncertainty distributions of our cost and QALY parameters. Further details on the distributions used for each input can be found in the Supplement, S1-S2. We than ran each of the 6,400 combinations of the model transition parameters, costs, and QALYs are then used to simulate 1,000,000 individuals to reduce the effect of first-order stochastic noise.[12]

#### Appendix Results

In our base-case results, we find that increasing the rate of deceased donor transplantation by 25% has an ICER of $8,100 per QALY gained compared to the status quo rate of transplantation. Smaller increases to the rate of transplantation are weakly dominated, but if we are unable to increase the rate of transplantation by 25%, it is important to quantify the impact of these smaller increases. From a societal perspective, any increases in the rate of transplantation are considered cost-saving. If we present cost-saving interventions as ICERs, they have negative values, which are indiscernible from interventions that are more costly and less effective. To overcome this, we can use a commonly used metric in cost-effectiveness research, incremental net monetary benefit (INMB).

$$INMB = \lambda \times \Delta\bar{E} - \Delta\bar{C}$$

where $\lambda$ is the willingness-to-pay (WTP) thershold, $\Delta\bar{E}$ is incremental difference in mean health effects, and $\Delta\bar{C}$ is the difference in mean costs. We use analysis of variance (ANOVA) to test for differences in INMB between scenarios and the Wilcoxon test when comparing INMB between two subgroups.

Using INMB, increasing the rate of transplantation to 25% results in the highest expected benefit, but all rates results positive INMB. This means that we should increase the rate of transplantation as much possible using imperfect but transplant quality kidneys. The INMB for increasing the rate of transplantation by 25% is $52,800 (95% CrI: $35,500-$80,800) and $66,100 (95% CrI: $47,700-$91,800) from the healthcare sector and societal perspectives, respectively (**Figure S8**). The expected INMB of increasing the rate of transplantation by 25% is somewhat smaller with older ages but substantially greater than 0 and hence remains the preferred strategy (**Figure S10**). Candidates with diabetes have lower, but still positive, expected INMB from both perspectives (**Figure S11**). We also find that compared to NH White candidates, all other race/ethnicity groups benefit more from a higher rate of transplantation (**Figure S12**).

#### Figure S4: Incremental cost-effectiveness frontier by age group and perspective.

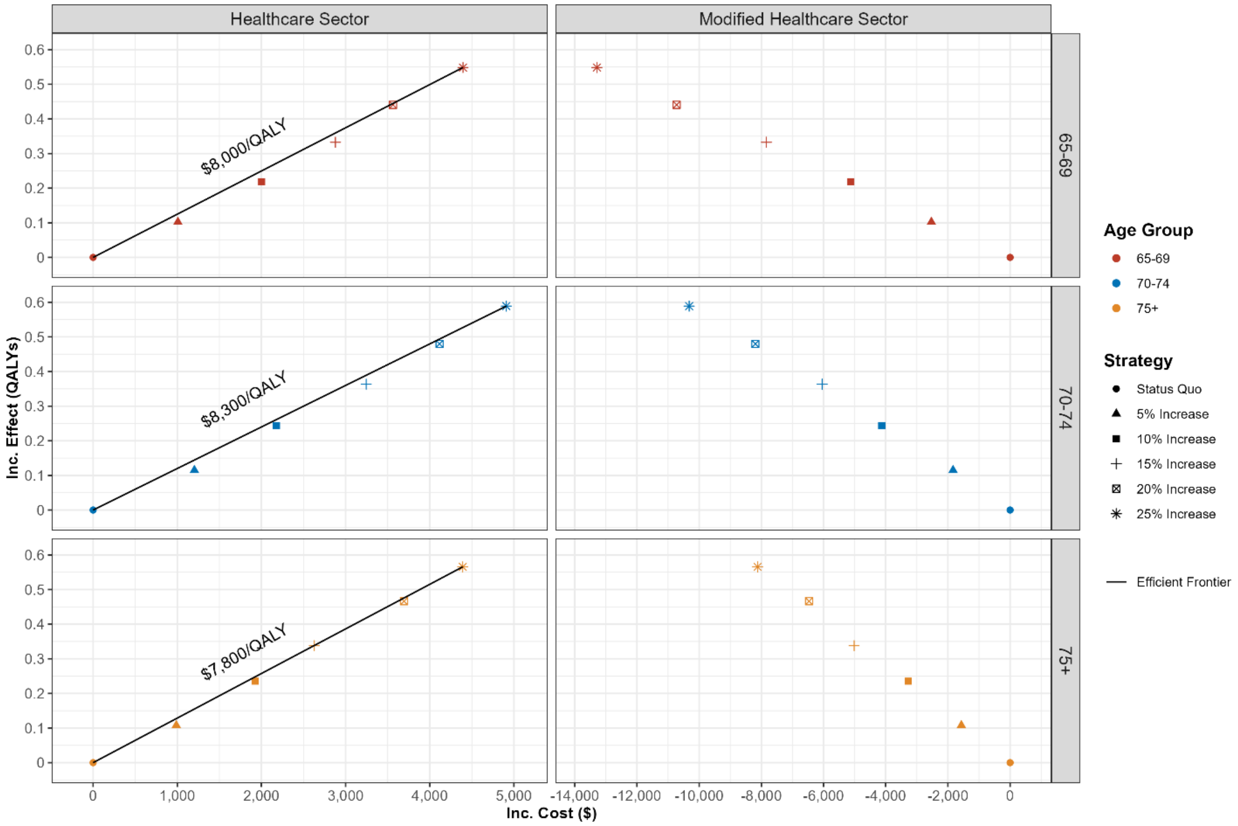


QALYs: quality-adjusted life years
Inc: Incremental

#### Figure S5: Incremental cost-effectiveness frontier by race and ethnicity group and perspective

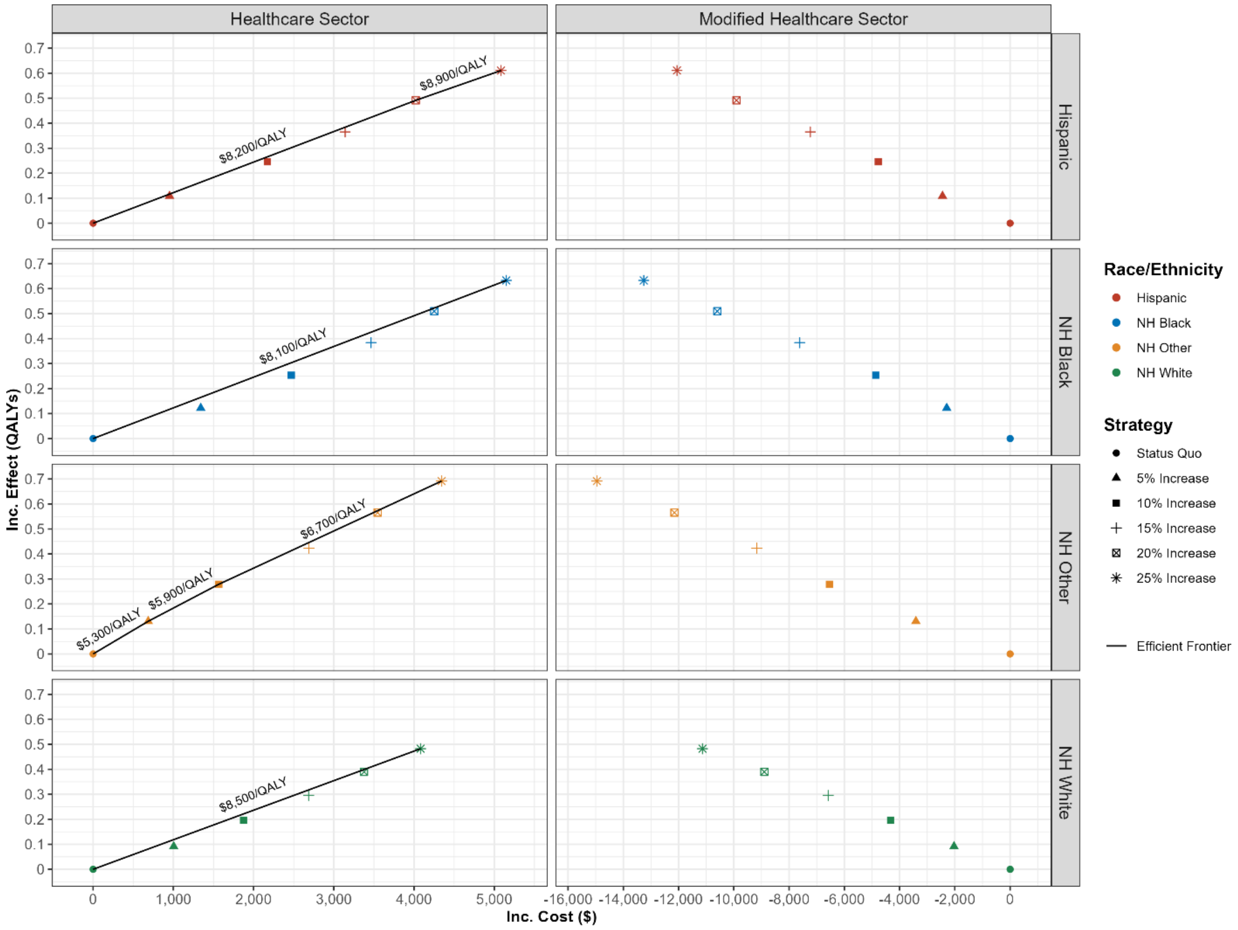


QALYs: quality-adjusted life years
Inc: Incremental

#### Figure S6: Incremental cost-effectiveness frontier by diabetes history and perspective

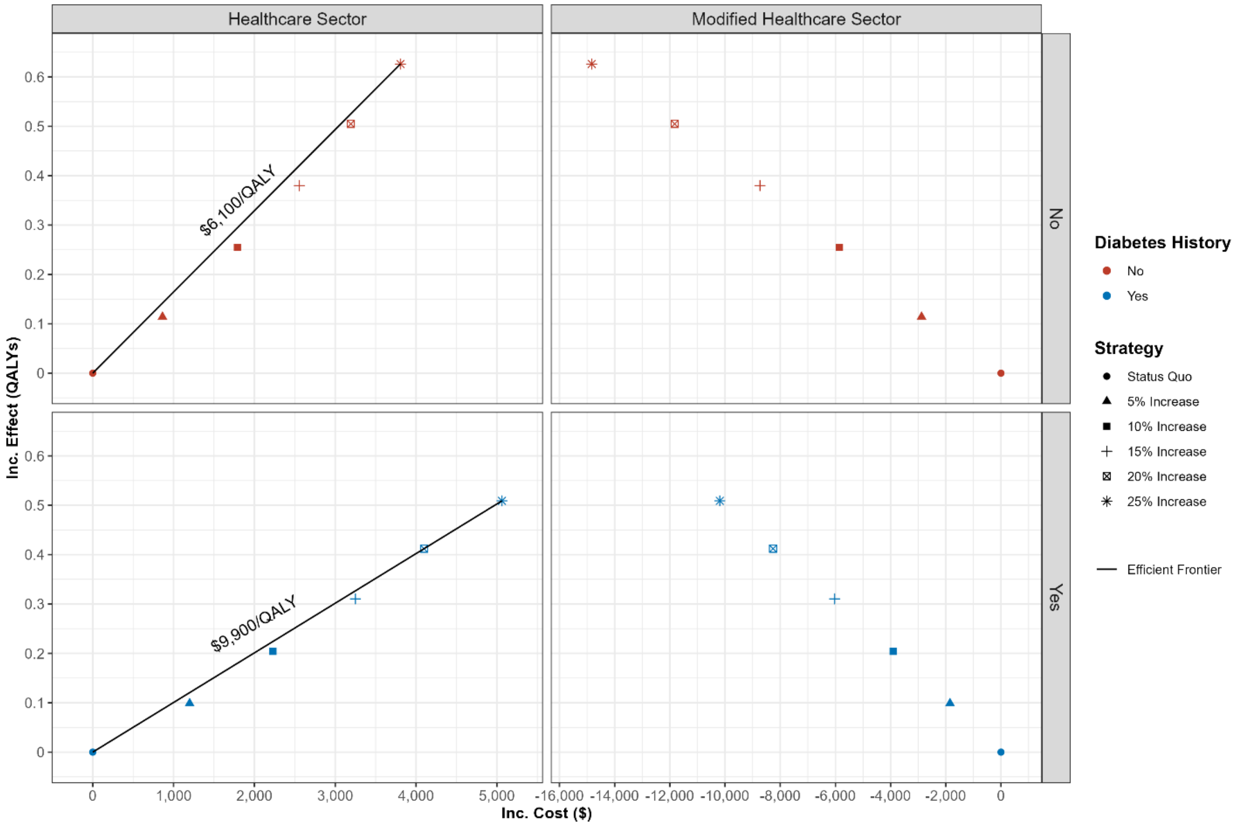


QALYs: quality-adjusted life years
Inc: Incremental

#### Figure S7: Incremental cost-effectiveness frontier by perspective and the percentage of kidneys with worse quality than their KDPI implies.

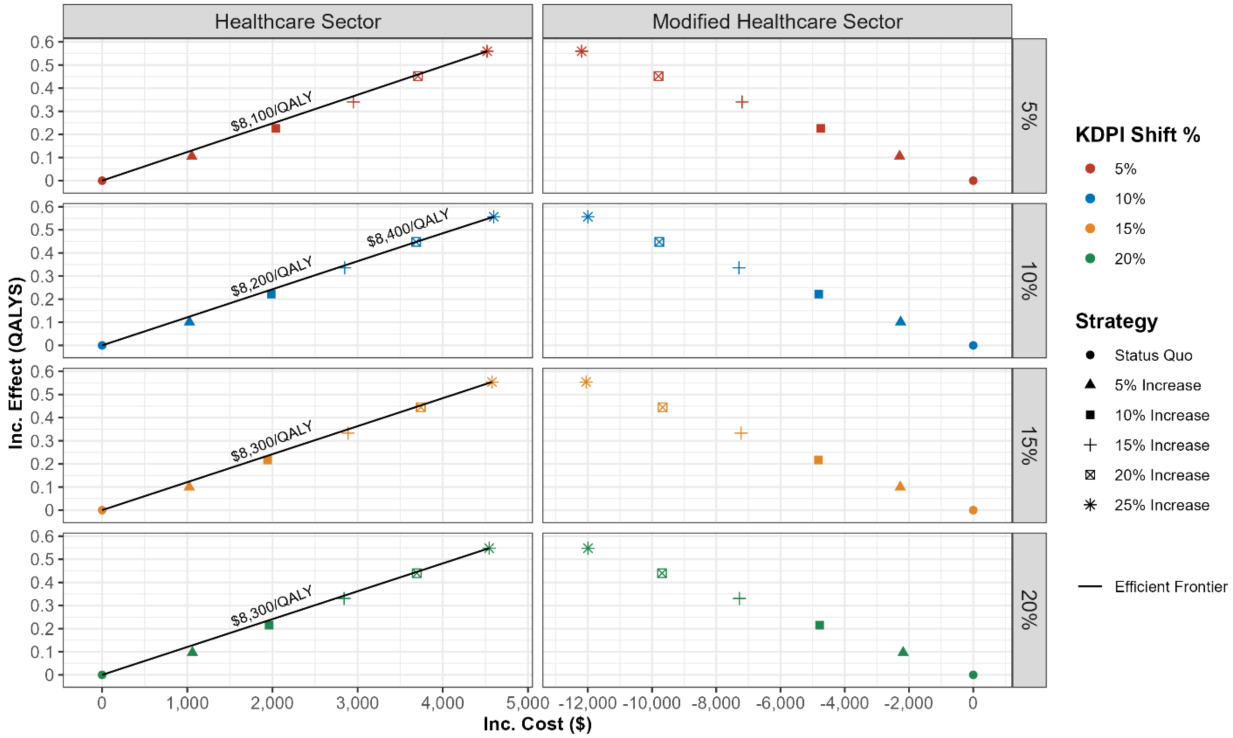


QALYs: quality-adjusted life years
Inc: Incremental
KDPI: kidney donor profile index

#### Figure S8: INMB by perspective

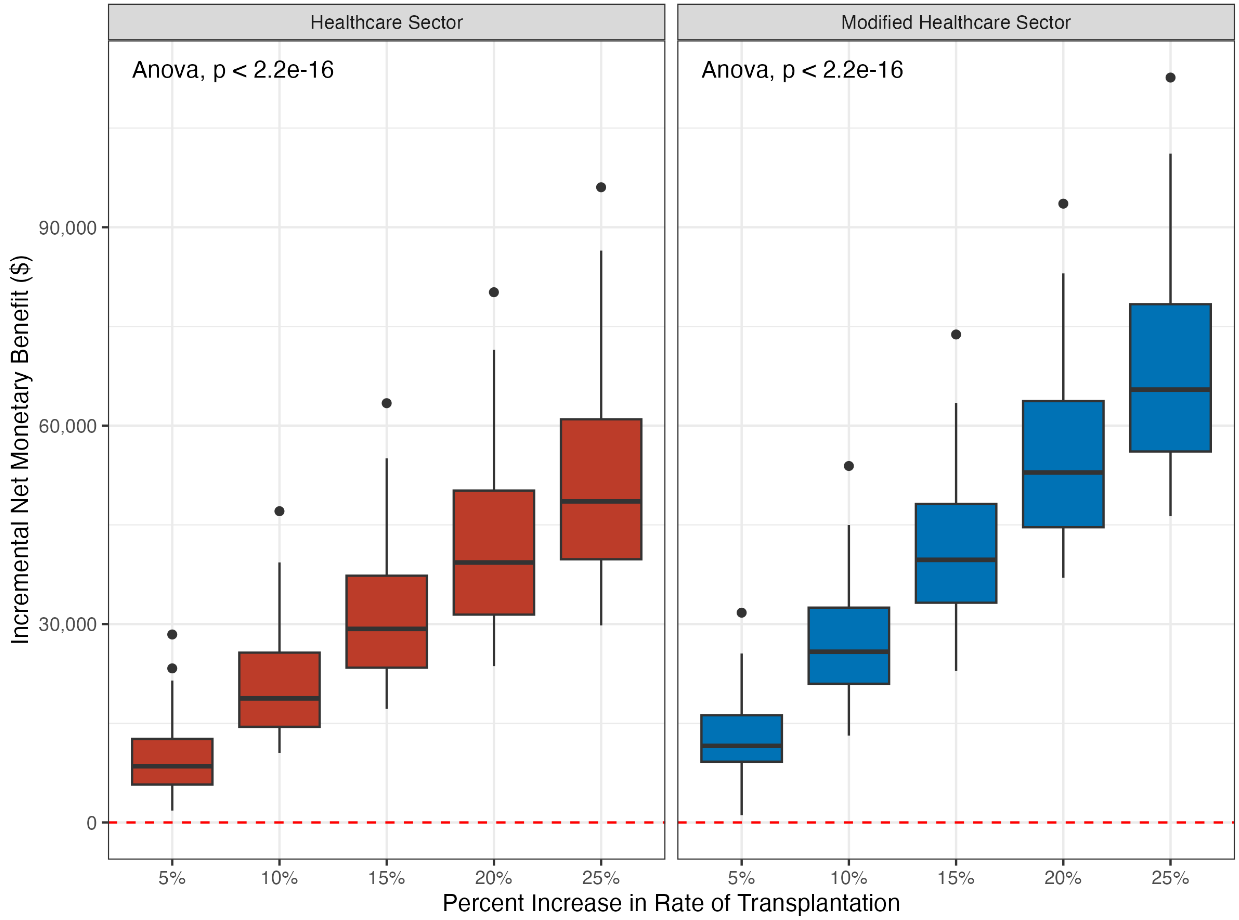


A comparison of means is tested using ANOVA tests, where the null hypothesis is that there is no difference in the mean INMB across the increasing rates of transplantation.
INMB: Incremental net monetary benefit

#### Figure S9: INMB by key subgroup

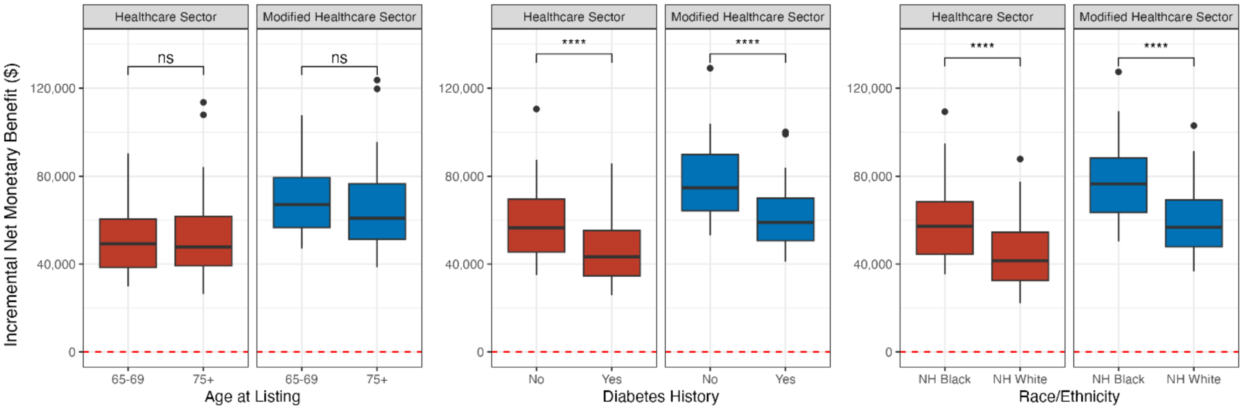


A comparison of means is tested using a Wilcoxon test, where the null hypothesis is that there is no difference in the mean INMB between the two subgroups. Statistical significance symbols: ns – $p > 0.05$, **** $p \le 0.0001$.
INMB: Incremental net monetary benefit

#### Figure S10: INMB by age group

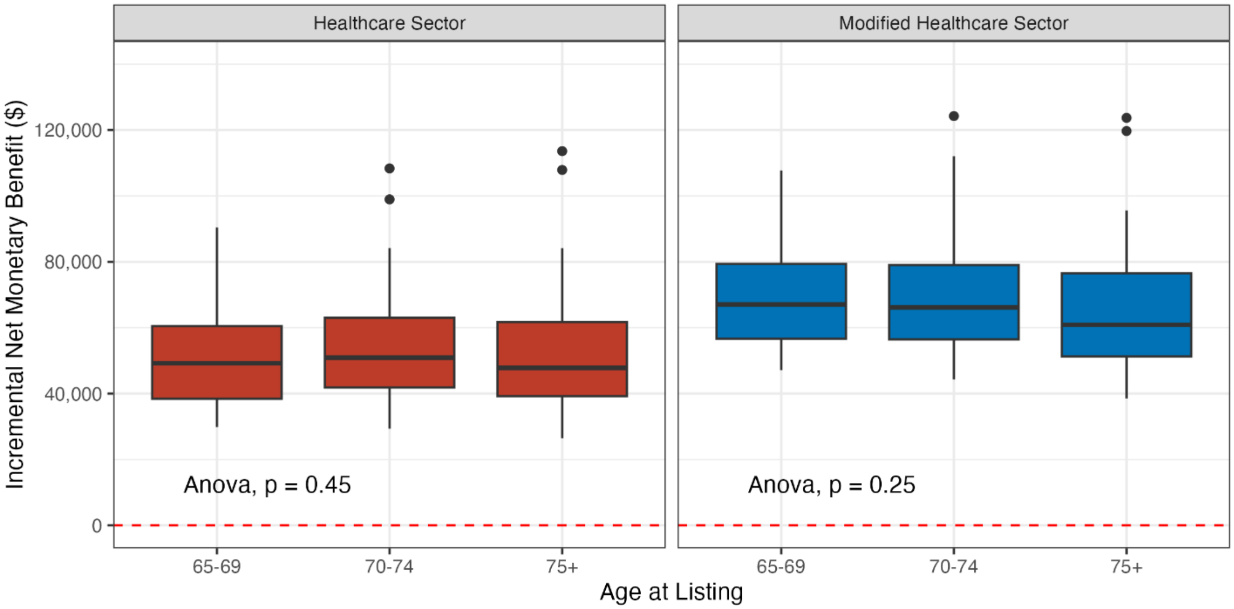


A comparison of means is tested using a ANOVA test, where the null hypothesis is that there is no difference in the mean INMB between the two subgroups. There is no statistically significant difference in INMB by age group.
INMB: Incremental net monetary benefit

#### Figure S11: INMB by diabetes status

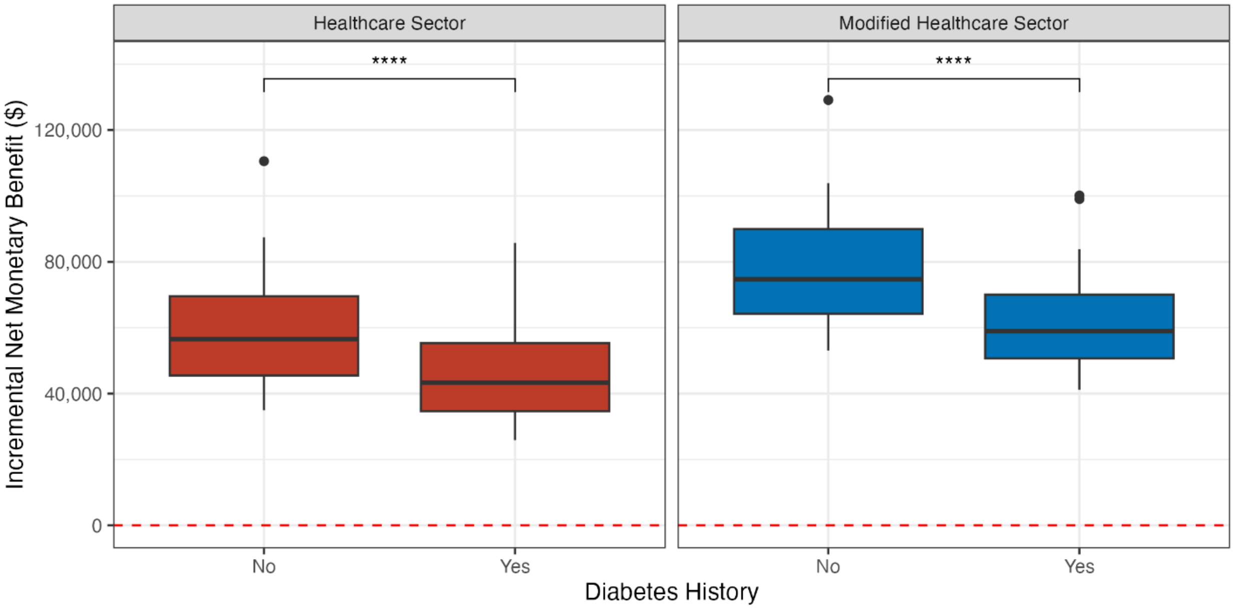


A comparison of means is tested using a Wilcoxon test, where the null hypothesis is that there is no difference in the mean INMB between the two subgroups. Statistical significance symbols: **** $p \leq 0.0001$.
INMB: Incremental net monetary benefit

#### Figure S12: INMB by race and ethnicity subgroup

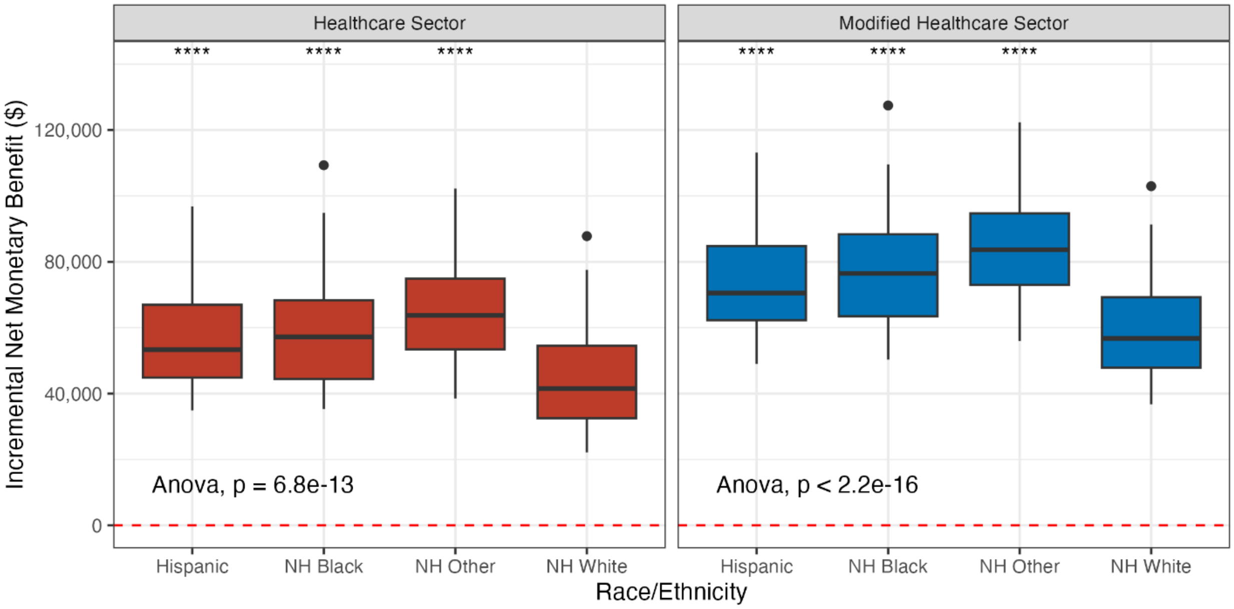


A comparison of means is tested using a Wilcoxon test, where the null hypothesis is that there is no difference in the mean INMB between NH White candidates and each other race/ethnicity group. Statistical significance symbols: **** $p \leq 0.0001$.

INMB: Incremental net monetary benefit

#### Figure S13: INMB for the percentage of kidneys with worse quality than their KDPI implies

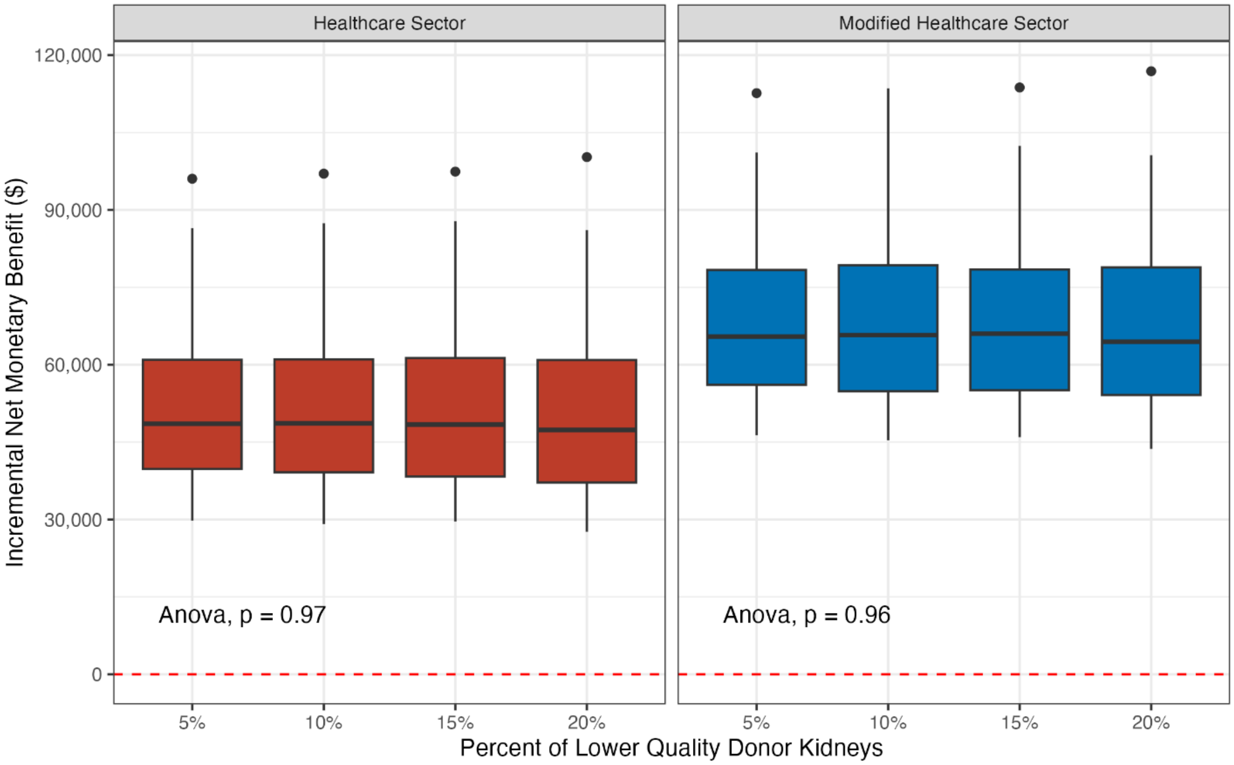


A comparison of means using ANOVA tests, where the null hypothesis is that there is no difference in the mean INMB across the percentage of kidneys with lower quality.
INMB: Incremental net monetary benefit

#### Figure S14: Expected loss curves by perspective

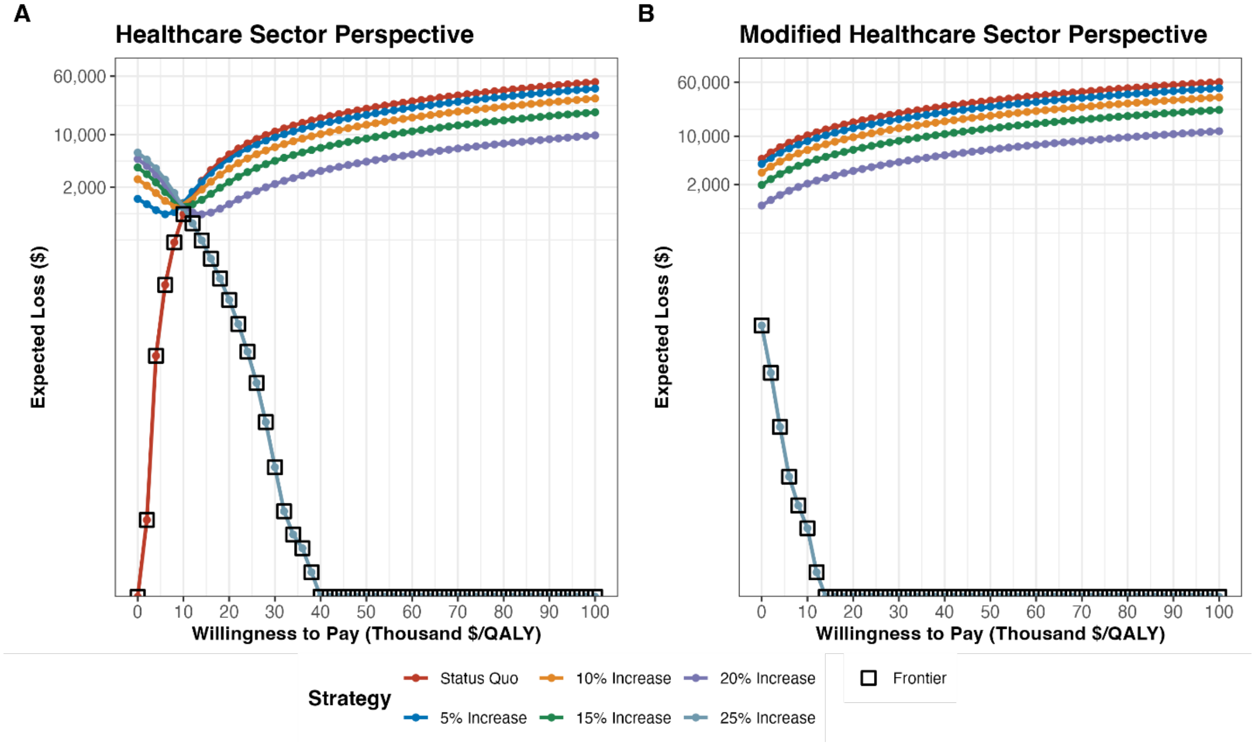


Expected loss curves from A) healthcare sector perspective, and B) modified healthcare sector perspective.
QALY: quality-adjusted life year
